# Characteristics and HIV-related Engagement of Male Sexual Partners of Female Sex Workers in Sub-Saharan Africa (SSA): a Scoping Review

**DOI:** 10.1101/2024.12.03.24318456

**Authors:** Galven Maringwa, Primrose Matambanadzo, James R. Hargreaves, Elizabeth Fearon, Frances M. Cowan

## Abstract

**Introduction:** Understanding the characteristics and behaviors of male sexual partners of female sex workers (FSWs) is crucial for comprehending the dynamics of HIV transmission. We aimed to explore and quantify the existing literature on male sexual partners of FSWs in SSA, where HIV prevalence is high and the dynamics of sex work are poorly understood. We focused on the proportions of men reporting sex with FSWs, along with their characteristics, HIV prevalence, and engagement with HIV services among the general population and specific subgroups.

**Methods:** We searched the literature in the EBSCOhost databases (Medline Complete, Global Health, and CINAHL). We included quantitative epidemiological peer-reviewed articles published in English between January 2010 and December 2023, following PRISMA guidelines for scoping reviews. The last search was performed on 09 October 2024. Eligible studies focused on men from the general population and subgroups of men who reported having sex with FSWs. The results were synthesized narratively to identify patterns and gaps in the literature.

**Results:** We identified 2,067 articles and reviewed 15, including one meta-analysis. The general population meta-analysis and the articles among subgroups revealed variations in reporting sex with FSWs, differences in HIV prevalence, suboptimal HIV testing uptake, and inconsistent condom use. The proportion of general population men in SSA who had ever paid for sex was 8.5%, with a pooled HIV prevalence of 3.6% and 67.5% reported condom use at last paid sex. High-risk subgroups of men reported different rates of sex with FSWs, ranging from 6.6% to 74%. HIV prevalence also varied significantly, from 7.5% to 26%. Across these high-risk groups, suboptimal HIV testing uptake and inconsistent condom use were common, with no comparative data for men who did not have sex with FSWs.

**Discussion:** Men who have sex with FSWs cannot be identified by specific characteristics. High-risk subgroups reported greater engagement with FSWs. Sex with an FSW was associated with higher HIV prevalence among men in the general population, with no data on subgroups. These findings highlight the need for tailored, occupation-specific interventions that address the unique needs of mobile and high-risk men.

## Introduction

Sub-Saharan Africa is disproportionately affected by HIV, accounting for nearly two-thirds (65%) of the global burden of infection. Eastern and Southern Africa alone is home to 52% of the global total(1). FSWs in southern Africa have a particularly high incidence and prevalence of HIV infection(2, 3). However, research on male sexual partners of FSWs is limited. Understanding the impact of male sexual partners of FSWs on sexual partnership dynamics, and patterns of HIV transmission across different regions is important[4, 5]. Partnerships involving male sexual partners of FSWs are diverse, ranging from intimate partners including spouses and boyfriends to casual partners who pay for sex. These partnerships may be built on love, trust, emotional intimacy, and financial need or dependency.

In general, men have poor uptake of HIV services or other healthcare services compared to women(4, 5). This poses challenges for providing effective prevention and care(6). Additionally, male sexual partners of FSWs do not form a visible and coherent social grouping which makes them difficult to target with interventions(7). Men who have sex with FSWs across SSA are at high risk of HIV. Certain subgroups of men have been associated with a higher prevalence of having sex with FSWs, potentially due to the nature of their work which involves mobility, and extended periods of separation from their spouses. These circumstances can enable and increase sexual interactions with FSWs(8, 9). These include men working in transportation and logistics, men in mining and extractive industries, uniformed service personnel, men in the informal sector, and seasonal migrant labourers. They have the potential to both transmit and acquire HIV from FSWs. Additionally, they can transmit HIV to their low-risk regular partners or spouses in the general population, potentially increasing the incidence of HIV in broader communities.

Studies on HIV epidemiology among men in these population subgroups conducted before 2010 found high rates of sex with FSWs and HIV prevalence among men in occupations closely associated with sex work. In 1991, among 331 truck drivers in East/Central Africa, HIV prevalence was 18%, with higher rates in Central Africa (31.75%) compared to East Africa (16.65%)(10).

A study conducted in 2002 among 480 Nigerian naval personnel, of whom 94.4% were male, found that 32.5% of male respondents had ever paid for sex, 19.9% had done so in the past 6 months, and 41% did not use condoms during recent paid sex(11). Another study conducted in South Africa in 2002 revealed that HIV prevalence among truckers and FSWs was 56%(8). In 2003, HIV prevalence was 13.4% among 526 Ivorian FSWs clients(12). Among 2,825 men surveyed in Kenya in 2008, 15% of sexually active men reported sex with FSWs(13).

## Research Objectives and Questions

This scoping review maps existing evidence on the prevalence of sexual interactions with FSWs, as well as the characteristics, behaviors, and HIV service uptake among men in the general population and other subgroups identified as having high HIV prevalence and high rates of interactions with FSWs since 2010. This information is important for informing future research directions and interventions specific to these populations in sub-Saharan Africa.

## Methods

### Overview

This review was guided by the Arksey and O’Malley (2005) methodological framework for scoping reviews comprising of the following five steps, (i) identify the research question, (ii) identify the relevant studies, (iii) study selection, (iv) charting the data, and (v) collating, summarizing and reporting data(14). The review was performed per Preferred Reporting Items for Systematic Reviews and Meta-Analyses extension for Scoping Reviews (PRISMA-ScR) guidelines.

**Protocol registration:** The protocol was registered on the Open Science Framework (OSF)

### Stage One: Identifying the Research Questions

The overall research question is “What is known about the Characteristics and HIV-related Engagement of Male Sexual Partners of Female Sex Workers in Sub-Saharan Africa?”

The specific review questions to be addressed are:

1. What proportion of men in sub-Saharan Africa have sex with FSWs?
2. What socio-demographic and behavioral characteristics define men who have sex with FSWs in sub-Saharan Africa?
3. How does HIV prevalence differ among men who have sex with FSWs in sub-Saharan Africa?
4. To what extent are men who have sex with FSWs in sub-Saharan Africa engaged in sexual health services?

### Stage Two: Identifying Relevant Studies

Peer-reviewed articles were searched from 3 electronic databases: EBSCOhost (Medline Complete, Global Health, and CINAHL). We developed search terms and their variations and then combined search terms using Boolean operators “OR” and “AND” using parentheses to group related terms and operators for clarity. For SSA, we searched for each of the countries in SSA. We also used the Medical Subject Headings (MeSH) for each search term – Table 1. To refine our search, we applied filters including publication date restrictions, language restrictions, study design, geography, source types, and age filters. The publication year was restricted to 2010 to 2023.

**TABLE 1:**
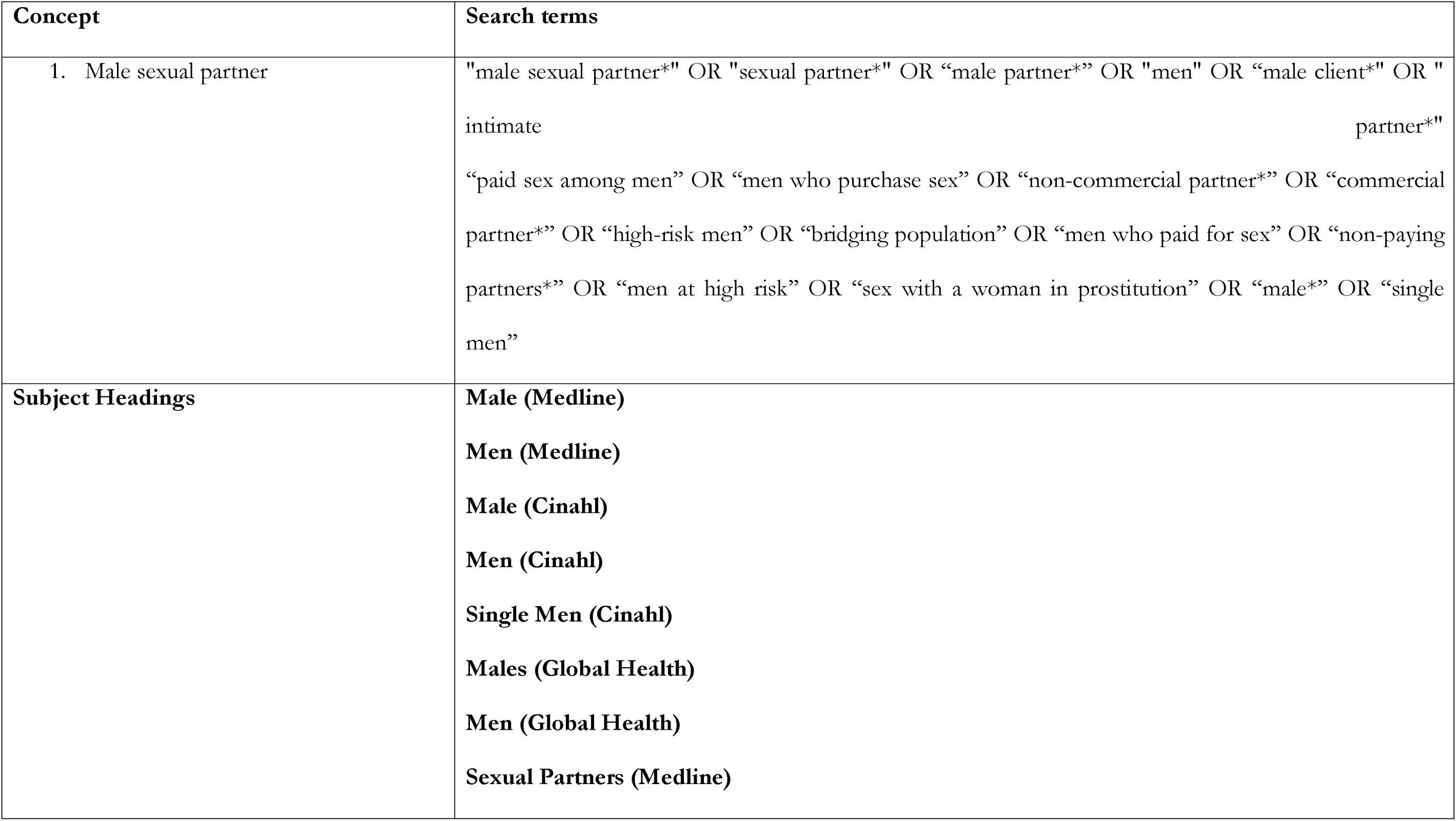
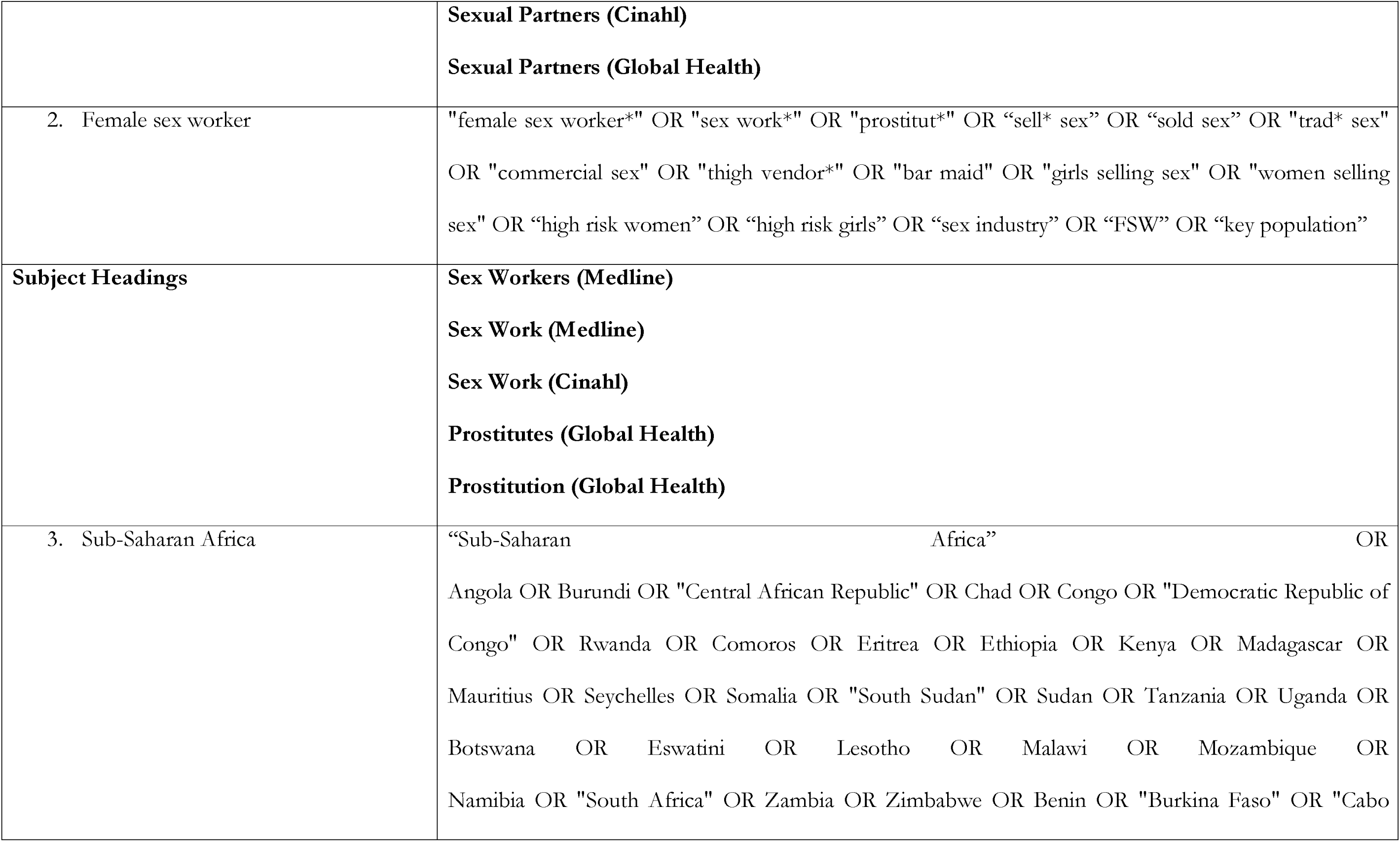
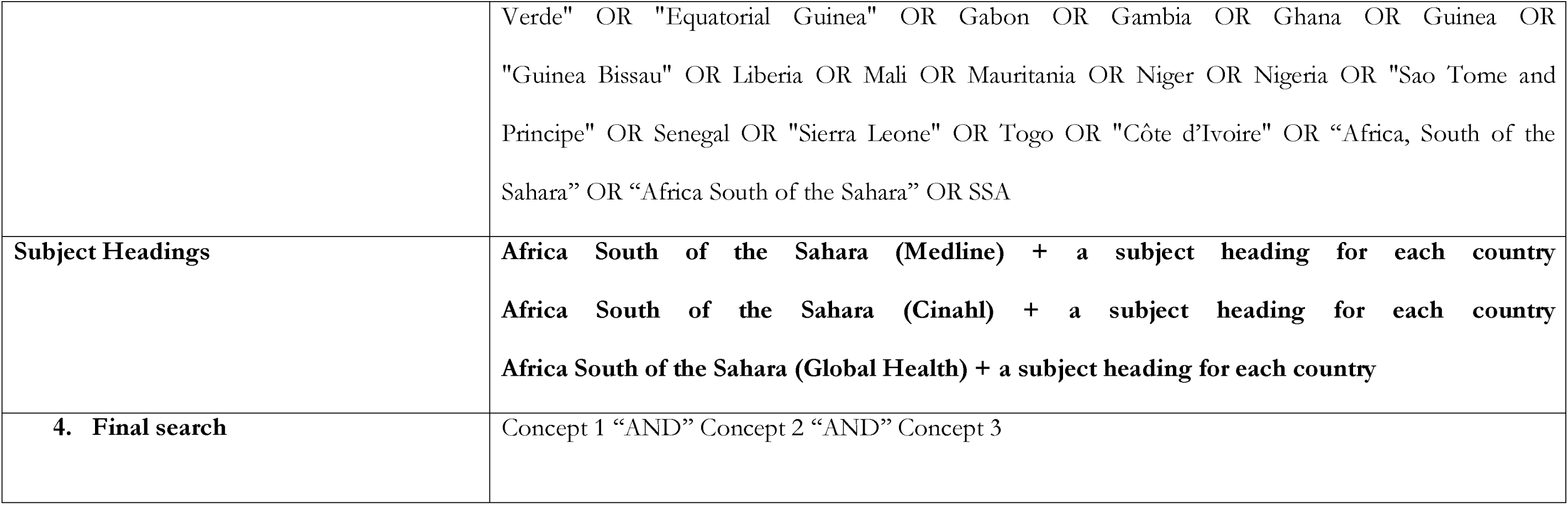
Concepts and search terms.

| Concept | Search terms |
| --- | --- |
| 1. Male sexual partner | "male sexual partner*" OR "sexual partner*" OR "male partner*" OR "men" OR "male client*" OR "intimate partner"<br>"paid sex among men" OR "men who purchase sex" OR "non-commercial partner*" OR "commercial partner*" OR "high-risk men" OR "bridging population" OR "men who paid for sex" OR "non-paying partners*" OR "men at high risk" OR "sex with a woman in prostitution" OR "male*" OR "single men" |
| Subject Headings | **Male (Medline)**<br>**Men (Medline)**<br>**Male (Cinahl)**<br>**Men (Cinahl)**<br>**Single Men (Cinahl)**<br>**Males (Global Health)**<br>**Men (Global Health)**<br>**Sexual Partners (Medline)** |
|  | **Sexual Partners (Cinahl)**<br>**Sexual Partners (Global Health)** |
| 2. Female sex worker | "female sex worker*" OR "sex work*" OR "prostitut*" OR “sell* sex” OR “sold sex” OR "trad* sex"<br>OR "commercial sex" OR "thigh vendor*" OR "bar maid" OR "girls selling sex" OR "women selling sex" OR “high risk women” OR “high risk girls” OR “sex industry” OR “FSW” OR “key population” |
| **Subject Headings** | **Sex Workers (Medline)**<br>**Sex Work (Medline)**<br>**Sex Work (Cinahl)**<br>**Prostitutes (Global Health)**<br>**Prostitution (Global Health)** |
| 3. Sub-Saharan Africa | “Sub-Saharan Africa” OR<br>Angola OR Burundi OR "Central African Republic" OR Chad OR Congo OR "Democratic Republic of Congo" OR Rwanda OR Comoros OR Eritrea OR Ethiopia OR Kenya OR Madagascar OR Mauritius OR Seychelles OR Somalia OR "South Sudan" OR Sudan OR Tanzania OR Uganda OR Botswana OR Eswatini OR Lesotho OR Malawi OR Mozambique OR Namibia OR "South Africa" OR Zambia OR Zimbabwe OR Benin OR "Burkina Faso" OR "Cabo |
|  | Verde" OR "Equatorial Guinea" OR Gabon OR Gambia OR Ghana OR Guinea OR "Guinea Bissau" OR Liberia OR Mali OR Mauritania OR Niger OR Nigeria OR "Sao Tome and Principe" OR Senegal OR "Sierra Leone" OR Togo OR "Côte d'Ivoire" OR “Africa, South of the Sahara” OR “Africa South of the Sahara” OR SSA |
| **Subject Headings** | **Africa South of the Sahara (Medline) + a subject heading for each country**<br>**Africa South of the Sahara (Cinahl) + a subject heading for each country**<br>**Africa South of the Sahara (Global Health) + a subject heading for each country** |
| **4. Final search** | Concept 1 “AND” Concept 2 “AND” Concept 3 |

### Stage Three: Study Selection

Studies were selected in three stages – retrieval, screening, and data extraction.

#### Article retrieval

In the first stage, GM used the above-mentioned strategy to search literature. All duplicate articles were removed using Endnote version 20 reference management software with a ‘find duplicates’ function. All articles were then exported to the Covidence reference manager for review. A trail of the electronic searches was saved on users’ EBSCOhost accounts: Medline Complete, CINAHL, and Global Health. Inappropriate articles were excluded at this stage.

#### Screening

In the second stage, a titles and abstracts review was conducted by GM, then a full-text review against the inclusion criteria. GM and PM independently conducted article screening for relevance and resolved all conflicts documenting reasons for article exclusions. The population in the synthesis comprises men from the general population and specific high-risk subgroups in SSA. These subgroups, such as truck drivers, miners, and military personnel, are characterized by high mobility. While the subgroups were not pre-specified, all the articles analyzed included questions about whether the men had had sex with an FSW or paid for sex. Full-text articles were retrieved after the reviewers reached a consensus on which articles to include based on predefined criteria. This ensured a comprehensive evaluation of the articles to determine their relevance and suitability for the research or review project. During the conflict resolution process, we encountered no significant discrepancies requiring formal reporting.

#### Data extraction

GM extracted data from the articles that met the inclusion criteria. The electronic search strategy retrieved 2,067 studies, from which 410 duplicates (19.8%) were removed. We screened the titles and abstracts of the 1657 (80.2%) that remained. Sixty-five (65) studies underwent full-text review, of which 15 (23.1%) met the inclusion criteria. Most of the relevant studies were retrieved from the Medline Complete database. The search results and the study inclusion process are reported in full and presented in a PRISMA-ScR flow diagram – Fig 1.

**Fig 1:**
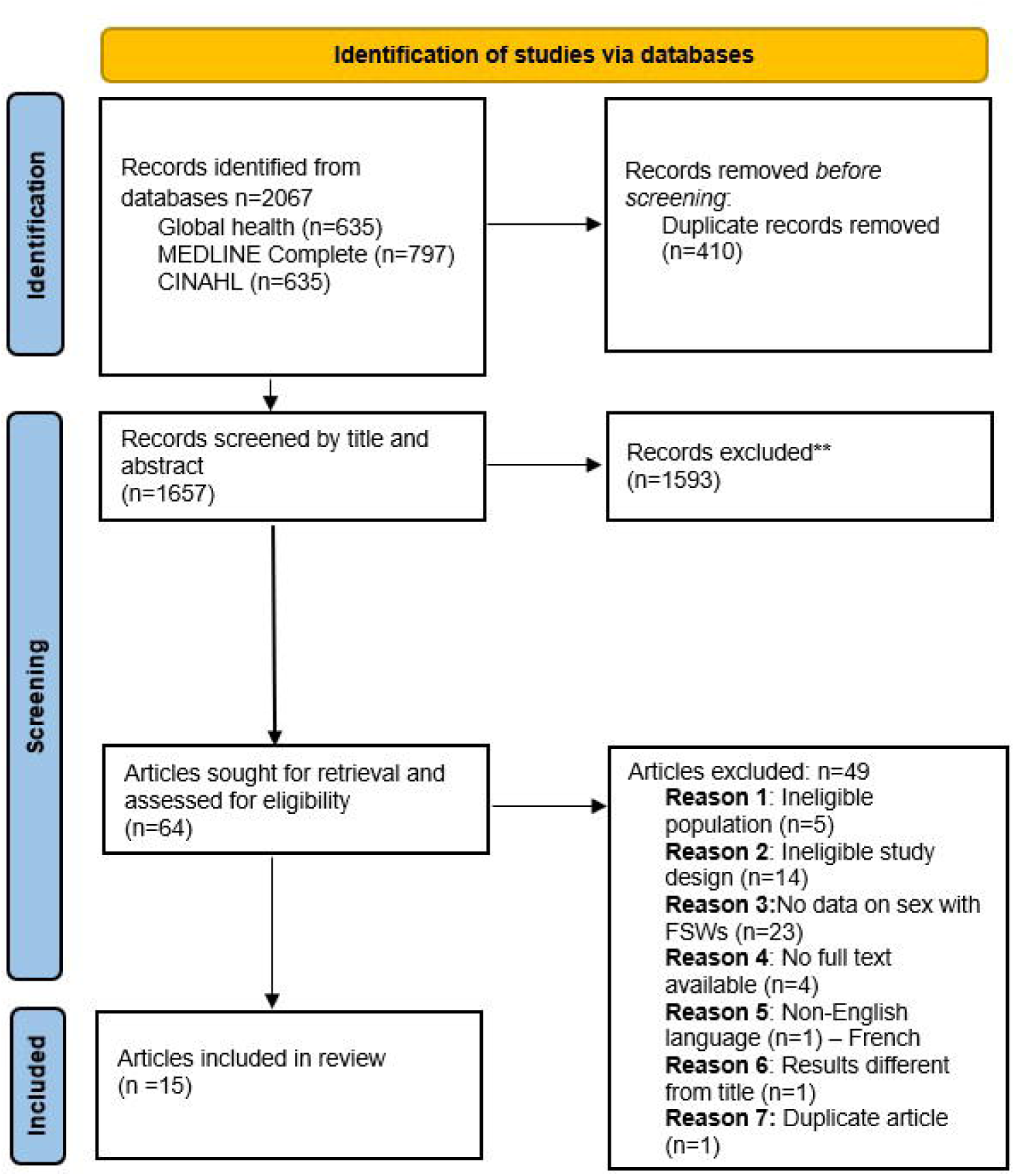
PRISMA_ScR Flow diagram of the study selection process

### Stage Four: Charting the Data

A single Excel spreadsheet was used to compile the study characteristics that were extracted from full articles for validation. Data collected from each study included: authors, country, population type, concept, study design, sampling strategy, survey dates, participants’ socio-demographics, sexual behaviour, and HIV prevalence. We also reported reasons for study exclusion for all excluded articles (S2 Appendix).

### Stage Five: Collating, Summarizing, and Reporting the Results

We conducted a narrative review aligned with the review objectives and questions, presenting the findings in graphs, tables, and narrative summaries.

#### Methodological quality appraisal and data synthesis

We selected the scoping review method to outline the different types of evidence related to our area of interest and identify gaps for further research. We used the Appraisal tool for Cross-Sectional Studies (AXIS) to appraise studies that we included for the review(15) – S1 Appendix.

#### Ethics and dissemination

We did not require ethical approvals as this was a literature review.

## Results

### Characteristics of the included studies

The surveys reported in the meta-analysis are distributed as follows: Central Africa (n=7, 10.9%), Western Africa (n=27, 42.2%), Eastern Africa (n=23, 35.9%), and Southern Africa (n=7, 10.9%). Surveys included in the metanalysis were the AIDS Indicator Survey (AIS), Demographic and Health Surveys (DHS), Population-based HIV Impact Assessment (PHIA), the Kenya AIDS Indicator Survey (KAIS), and the South African National HIV Prevalence, Incidence, Behaviour and Communication Survey (SABSSM). These studies were used to assess population sizes, HIV prevalence, and HIV prevention among men who paid for sex in SSA (2010–2020).

There were 14 studies among population subgroups included in the synthesis, of which 6 (42.8%) were conducted in the transport and logistics industry(17–21, 30), 3 (21.4%) among male patrons recruited in social venues (25–27), 2(14.3%) among personnel in the mining and extractive industry(23, 31), 2 (14.3%) among migrants and refugees(22, 23), and one among military personnel(32). These studies assessed varying outcomes including HIV prevalence, HIV testing history, condom use, and paid sex prevalence. An overview of each of these studies is shown in Table 2.

**TABLE 2:** SUMMARY OF EACH REVIEWED STUDY.

|  | **Authorship** | **Country** | **Population/**<br>**Participants** | **Concept** | **Participant**<br>**characteristics** | **Study Design** | **Sampling strategy** | **Dates of**<br>**survey** |
| --- | --- | --- | --- | --- | --- | --- | --- | --- |
| 1. | W. Gaolalwe,<br>et al., 2023(16) | Botswana | Long-distance<br>truck drivers<br>(LDTD). | Knowledge of HIV<br>transmission ways,<br>uptake of HIV<br>prevention strategies eg<br>condoms | Age, religion,<br>education, age at start<br>of driving, duration in<br>the industry,<br>relationship status | Cross-sectional | Simple random<br>sampling technique | February to<br>August 2017. |
| 2. | Adeoti,<br>Adekunle<br>Olatayo, et al.,<br>2021(17) | Nigeria | Long-distance<br>truck drivers<br>(LDTD). | Perceived sexual risk<br>factors and willingness<br>to undergo HIVST. | Age, duration as a<br>driver, BMI, smoking<br>history, alcohol<br>consumption, paid sex,<br>age at first sex, number<br>of partners, drug use | Cross-sectional | Systematic random<br>sampling | Sept – Dec<br>2019 |
| 3. | <u>Sinead Delany-</u><br><u>Moretlwe</u>,<br>2013 | South<br>Africa | Long-distance<br>truck drivers<br>(LDTD). | Prevalence of HIV and<br>correlates of HIV<br>infection | Age, marital status,<br>province of residence,<br>education, income, job<br>type, access to medical<br>treatment, language,<br>years as a trucker, time on the road, frequency away from home, have another home, knowledge of STI, awareness of truckers' clinics, knowledge of HCT, perceived HIV risk, testing history. | Cross-sectional | Two-stage stratified<br>sampling<br>Stage 1: A 10%<br>random sample of<br>depots<br>was drawn from the list, stratified by small, medium, and large depots<br>Stage 2: All eligible drivers were recruited within each strata until sample size was reached | October<br>2003 and July<br>2004. |
| 4. | <u>Samanta Tresha Lalla-Edward,</u><br>2019(18) | South Africa | Long-distance truck drivers (LDTD). | Health status | Age, years driving, hours spent driving, days spent driving, country of origin, race, education, marital status, | Cross-sectional | Convenience sampling | October 2016 and March 2017 |
| 5. | Carlos Botão, | Mozambiq | Long-distance | HIV prevalence and | Age, language. | Cross-sectional | Convenience | February 23 |
|  | 2016(19) | ue | truck drivers (LDTD). | associated risk factors | Education, circumcision, marital status, nationality, number of trips per month |  | sampling | and August 26, 2012 |
| 6. | Yihunie Lakew, 2014(20) | Ethiopia | Taxi drivers and assistants | HIV related risk behaviours | Age, education, marital status, religion, duration as taxi driver, taxi ownership | Cross-sectional | Multistage cluster sampling<br>Stage 1: available clusters were identified based on the usual taxi density.<br>Stage 2: Random selection of participant from each cluster | April 8–17, 2006. |
| 7. | Christina P | Uganda | Motorcycle | Prevalence of HIV/ | Age | Respondent- | Respondent-driven | 2008 to 2009 |
|  | Lindan,<br>2015(21) |  | taxi drivers | sexually transmitted<br>infections | Years in school<br>Religion<br>Marital status<br>Circumcision<br>Number of partners<br>Condom use<br>HIV testing history<br>HIV status | driven sampling | sampling |  |
| 8. | Cynthia Sema'<br>Baltazar,<br>2014(22) | South<br>Africa | Miners | HIV Prevalence and<br>Risk Behaviors | Age, language, religion,<br>circumcision status,<br>areas of residence in<br>Mozambique and<br>South Africa,<br>education, occupation,<br>years worked in mines, | Cross-sectional | Time-location<br>sampling | Feb to<br>May 2012 |
| 9. | Hordofa | Ethiopia | Gold miners | HIV preventive | Age, religion, ethnicity, | Cross-sectional | Simple random | Jan 15, to Feb |
|  | Gutema<br>Abdissa,<br>2014(23) |  |  | behavior and<br>associated factors-<br>actions and practices<br>that individuals adopt<br>to reduce their risk of<br>acquiring HIV<br>infection | marital status,<br>education, monthly<br>income |  | sampling | 15, 2012. |
| 10 | Bonnie Robin<br>Tran, 2019(24) | Democratic<br>Republic<br>of the<br>Congo | Military<br>personnel | Relationship between<br>alcohol use and sexual<br>risk behaviors<br>including sex with<br>FSW | Age, education, marital<br>status, military rank,<br>military branch and<br>AUDIT | Cross-sectional | Multi-stage sampling<br>Convenience<br>sampling | October 2013<br>to April 2014 |
| 11 | Eric Abella<br>Roth, 2014(25) | Kenya | Male patrons<br>in venues –<br>PLACE<br>surveys | Investigating the<br>reasons, motivations,<br>and justifications<br>behind men's involvement in commercial sex within the specific cultural and economic context of the informal settlement | Age, marital status,<br>education, weekly<br>income, number of<br>children | Cross-sectional | Venue-based<br>sampling | July-December<br>2010 |
| 12 | Olanrewaju Olaniyan, 2017(26) | Nigeria | Male patrons in venues – PLACE surveys | Level of desire or request for heterosexual commercial sex services | Age, marital status, education, age at first sex, age at first paid sex debut, number of dependents, place of patronising, amount paid | Cross-sectional | Multi-stage sampling<br>Purposive sampling then random sampling | Not specified |
| 13 | Paul J. Fleming, 2015(27) | Ghana<br>Tanzania | Male patrons in venues | Role of relationship types on condom use | Age, salary in the past month, and education), attitudes towards | Time-space sampling<br>Cross-sectional | Time-space sampling | June to November 2008 in Tema, |
|  |  |  |  |  | gender equality, and self-reported sexually transmitted infections (STI) |  |  | Ghana<br>September 2008 to February 2009<br>in Mbeya, Tanzania |
| 14 | Kassa Tiruneh, 2015(28) | Ethiopia | Seasonal migrant laborers | Sexual behavior and vulnerability to HIV infection | Age<br>Religion<br>Ethnicity<br>Marital status<br>Education | Cross-sectional | Multistage random sampling<br>Stage 1: Random sampling of primary sampling units<br>Stage 2: systematic random sampling of participant per unit | July 8–18, 2013 |
| 15 | Caroline Hodgins(29) | SSA | Sexually active men | Population size, HIV prevalence, and use | Age<br>Residency | Multi-stage cluster | Comprehensive sampling | 2010 - 2020 |
|  |  |  | recruited to<br>population<br>representative<br>household<br>surveys in 34<br>countries | of HIV prevention and<br>treatment. | Population size<br>Lifetime number of<br>sexual partners<br>Condom use<br>HIV prevalence<br>HIV testing,<br>Antiretroviral<br>(ARV) use<br>Viral load suppression<br>(VLS) | sampling |  |  |

### Description of studies

A total of 15 studies were included in the synthesis, comprising 78 surveys. Of these, 64 were population-based (from 1 meta-analysis), 1 was a single study conducted in two countries (Ghana and Tanzania), and 13 were single-setting studies focused on sub-groups of men. These surveys were distributed as follows: 8 (10.3%) in 6 Central African countries, 30 (38.5%) in 13 Western African countries, 29 (37.2%) in 12 Eastern African countries, and 11 (14.1%) in 5 Southern African countries – Fig 2.

**Fig 2:**
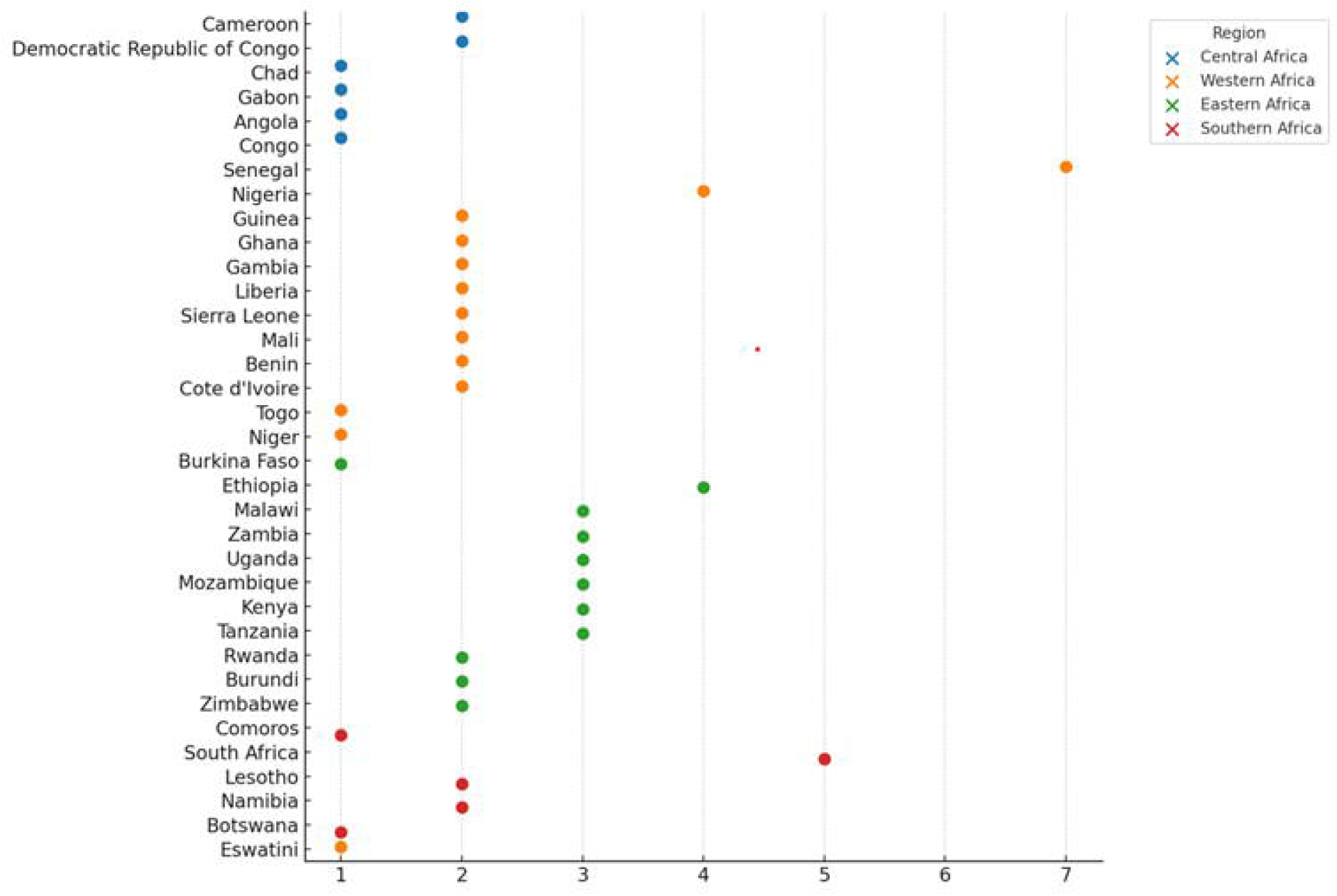
Number of studies by country and region

### Type of methods and design

All surveys included in the meta-analysis employed multi-stage cluster sampling to ensure national representativeness. Simple random sampling was used in two studies(23, 33). The sampling frames were drawn from all the LDTDs registered for services at the weighbridge, as well as from lists generated at the small mining sites where they worked. Systematic random sampling was utilized among LDTD in one study where all available truck drivers were numbered, and the truckers with odd numbers were selected(17). Multi-stage stratified sampling was employed in studies involving various groups: taxi drivers (from taxi stations to drivers), LDTDs (from depots to drivers), military personnel (from military bases to units, then soldiers), male patrons (from areas to brothels, then clients), and seasonal migrant labourers (from study sites to migrants)(20, 26, 28, 30, 32). Convenience sampling was employed among LDTD in two studies(18, 19). Time allocation sampling was used among miners and male patrons from social venues(25, 27, 31). Lastly, one study utilized respondent-driven sampling (RDS) among high-risk motorcycle taxi drivers.

### Sample sizes

The meta-analysis recruited a substantial proportion of men from 34 countries representing 95% of men in the SSA(29)The total number of men recruited in subgroups was 10,286. Fig 3 provides key findings across articles about sample size, and HIV prevalence by subgroup.

**Fig 3:**
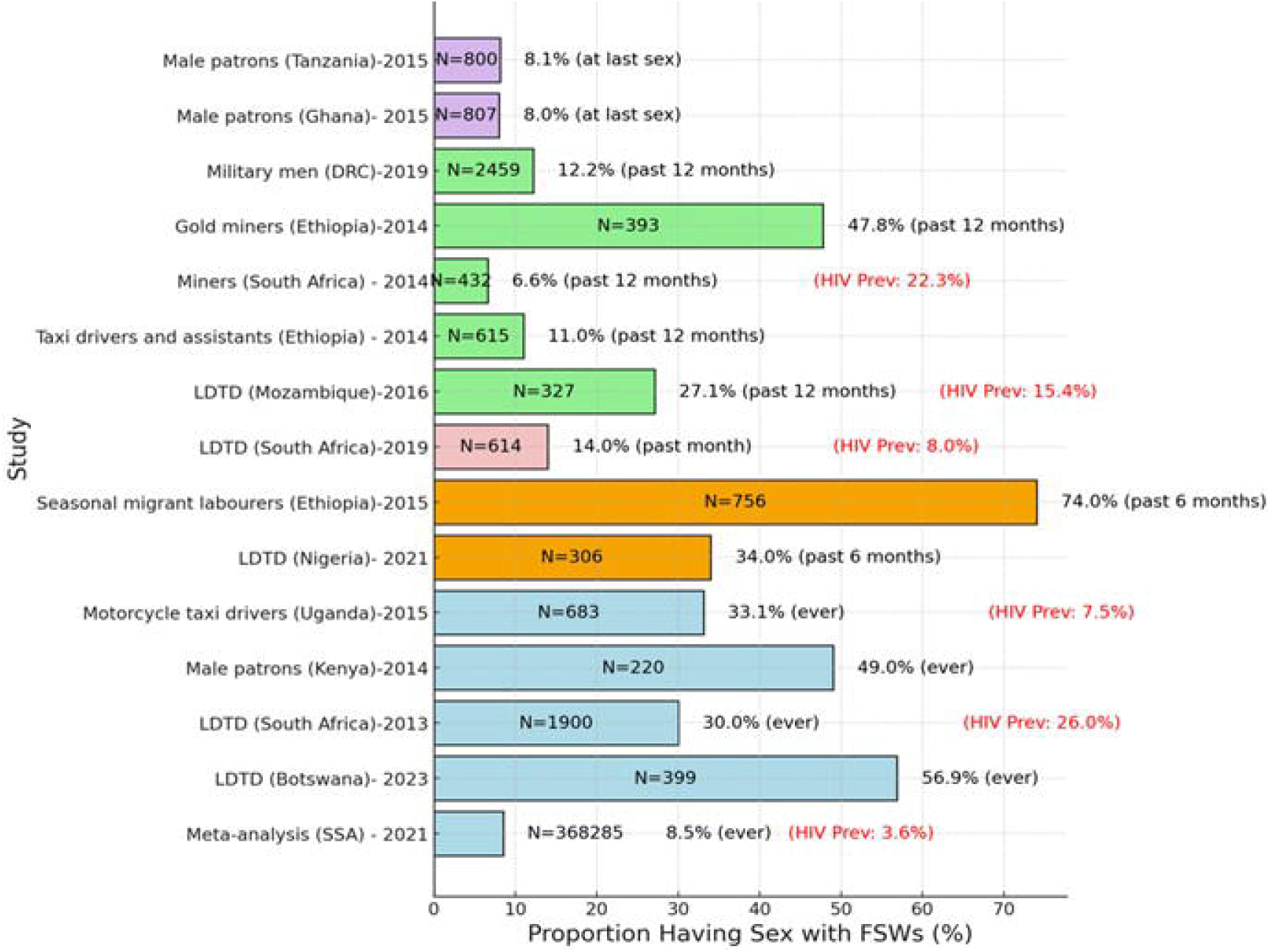
Sample size, proportion of men engaging with FSWs, and HIV prevalence

### Socio-demographic and behavioral characteristics of sexual partners of FSWs

The demographic characteristics of the research participants were not fully reported across all the studies. However, the meta-analysis included men aged 15-54 years, from both urban and rural areas(29). The socio-demographic characteristics presented for the subgroups apply to all men recruited in the studies. For the subpopulation studies, the age range spanned from 15 years among taxi drivers, assistants, and miners in Ethiopia, to over 50 years among LDTD in Nigeria, military men in DRC, male patrons in Kenya and Nigeria, and miners in South Africa(17, 20, 23–26, 31).

Marital and relationship status varied across studies. The proportion of married men ranged from 16.1% among social venue patrons to 88.5% among LDTD. Conversely, the rates of being single or unmarried ranged from 3.1% among LDTD in South Africa to 75.6% among taxi drivers and their assistants in Ethiopia. Other relationship types reported included steady girlfriends (22.3%) and casual girlfriends (9.5%) among LDTD in Botswana, 35.6% categorized as ‘partnered’ among migrants and refugees, and 3.6% who were ‘living together’ with sexual partners among LDTD in South Africa. Marital status was positively associated with HIV prevalence. Married men who were not staying with their spouses, as well as unmarried men who were staying with partners, were more likely to be HIV positive.

Depending on the country and context, different education categories were used with a lack of standardization across studies. Among military personnel in the DRC, 50.7% had completed primary education. In contrast, 36.0% of seasonal migrants in Ethiopia were unable to read or write. In Kenya, 42.0% of male patrons had only completed primary education, while 36.8% of high-risk motorcycle taxi drivers in Uganda had received 12 or more years of education. Men with primary education or lower were found to have higher odds of living with HIV among LDTD in Mozambique.

Alcohol and drug use varied by type of population. Alcohol use ranged between 64.0% among seasonal migrant labourers to 78.1% among LDTD in the past 6 months. Alcohol consumption was associated with higher HIV prevalence among seasonal migrant labourers.

### The proportion of men who pay for sex

The prevalence of paid sex varied considerably among studies in sub-Saharan Africa (SSA) mainly due to differences in study population and methodology.

#### Findings from the meta-analysis of the general population

The meta-analysis reported that 8.5% (95% CI: 6.4%-11.2%) of men in the general population self-reported ever paying for sex between 2010 and 2020[12]-Fig 3. The highest proportions were reported in Central and Eastern Africa at 15.2% (95% CI: 5.8-34.3) and 12.4% (95% CI: 8.3-18.0) compared to Western and Southern Africa at 6.0% (95% CI: 4.4-8.0), and 5.0% (95% CI: 2.7-9.1) respectively.

#### Findings from the high-risk subpopulation studies

The proportion of high-risk men who ever paid for sex ranged from 33.1% to 56.9%(16, 21). Among those who reported recent paid sex, the proportions ranged from 14% to 34-74% over the past 6 months(17, 18, 28). Paid sex in the past 12 months ranged from 6.6% to 27.1% highlighting high levels of heterogeneity in the prevalence of paid sex by subgroup and time variations(19, 31) – Fig 3.

#### Factors associated with having sex with an FSW

The meta-analysis did not report factors associated with paid sex. Only 6 articles among subgroups investigated factors associated with paid sex. In Botswana, LDTDs who had at least one girlfriend, were international rather than local drivers, had more years in the trucking industry, and lived with a wife rather than a girlfriend or alone or with family were more likely to have paid for sex in the past 12 months(33). Having multiple sexual partners in the past 6 months was associated with paid sex among LDTD in Nigeria(17). In Kenya, beliefs that men should practice sex before marriage, friends thinking that it is always right to have sex with FSWs in the bar, and that when they go to bars they always have enough money to afford an FSW were positively associated with having had sex with an FSW they met in a bar among male patrons recruited at public venues(25). Among military personnel in the DRC, probable problematic alcohol use was associated with having sex with an FSW(32). A study among male patrons in social venues in Nigeria found that younger men (<30 years), single or married but not living with their spouse, unemployed, earning more than 35,000 Naira, and patronizing street-based sex workers rather than brothels or houses were more likely to have sex with an FSW weekly(26).

### HIV prevalence

#### Findings from the meta-analysis of the general population

In the meta-analysis, men who reported having ever paid for sex were more likely to be HIV positive with a prevalence ratio of 1.50 (95% CI 1.31–1.72) compared to men who did not. The meta-analysis did not provide data on regional differences in the relationship between paying for sex and HIV prevalence(29). Across the studies, the pooled HIV prevalence was 3.6%, (95% CI 1.6% – 8.1%) in studies conducted between 2010 and 2020 – Fig 3.

#### Findings from the high-risk subpopulation studies

Only 5 studies among subgroups of men reported on HIV prevalence, and the data are not disaggregated by whether or not they paid for sex. HIV prevalence among LDTD in South Africa ranged from 7.1% (95% CI: 5.3-9.5) in 2019 to 26% (95% CI: 24.0-28.0) in 2013(18, 30). HIV prevalence was 22.3% (95% CI: 17.8-26.9) among miners in South Africa in 2014(31), 15.4% among LDTD in Mozambique in 2016, and 7.5% (95% CI: 5.2-10.0) among high-risk motorcycle taxi drivers in Uganda in 2015(21) – Fig 3.

#### Factors associated with HIV prevalence

In the meta-analysis, men who reported sex with FSWs were more likely to be living with HIV compared to those who did not. The meta-analysis did not examine any factors associated with HIV prevalence among men who had sex with FSWs. Four studies in subpopulations examined factors associated with HIV prevalence but did not compare men who had sex with FSWs and those who did not(19, 21, 30, 31). Many high-risk men were unaware of their HIV-positive status due to a lack of prior testing. Having a previous testing history was associated with 30% lower odds of being HIV-positive among LDTD in South Africa, compared to those with no prior HIV testing history(30). HIV among all men from high-risk groups was associated with factors such as being married, older age, low education, alcohol use, higher income, lack of HIV testing, living conditions allowing multiple partnerships, younger entry into truck driving, more time on the road, genital cleaning before sex, receiving healthcare abroad, foreign residency, and primary language. Limited HIV knowledge also correlated with higher prevalence in men frequently having sex with FSWs.

### Engagement in HIV prevention and care services

#### Condom use

##### Findings from the meta-analysis of the general population

The proportion of general population men who self-reported using a condom during their most recent paid sex was 67.5% (95% CI: 63.9%–70.9%)– Fig 4. Condom use data was not reported for men who did not have sex with FSWs. The meta-analysis did not disaggregate condom use proportions by region for the same period(29).

**Fig 4:**
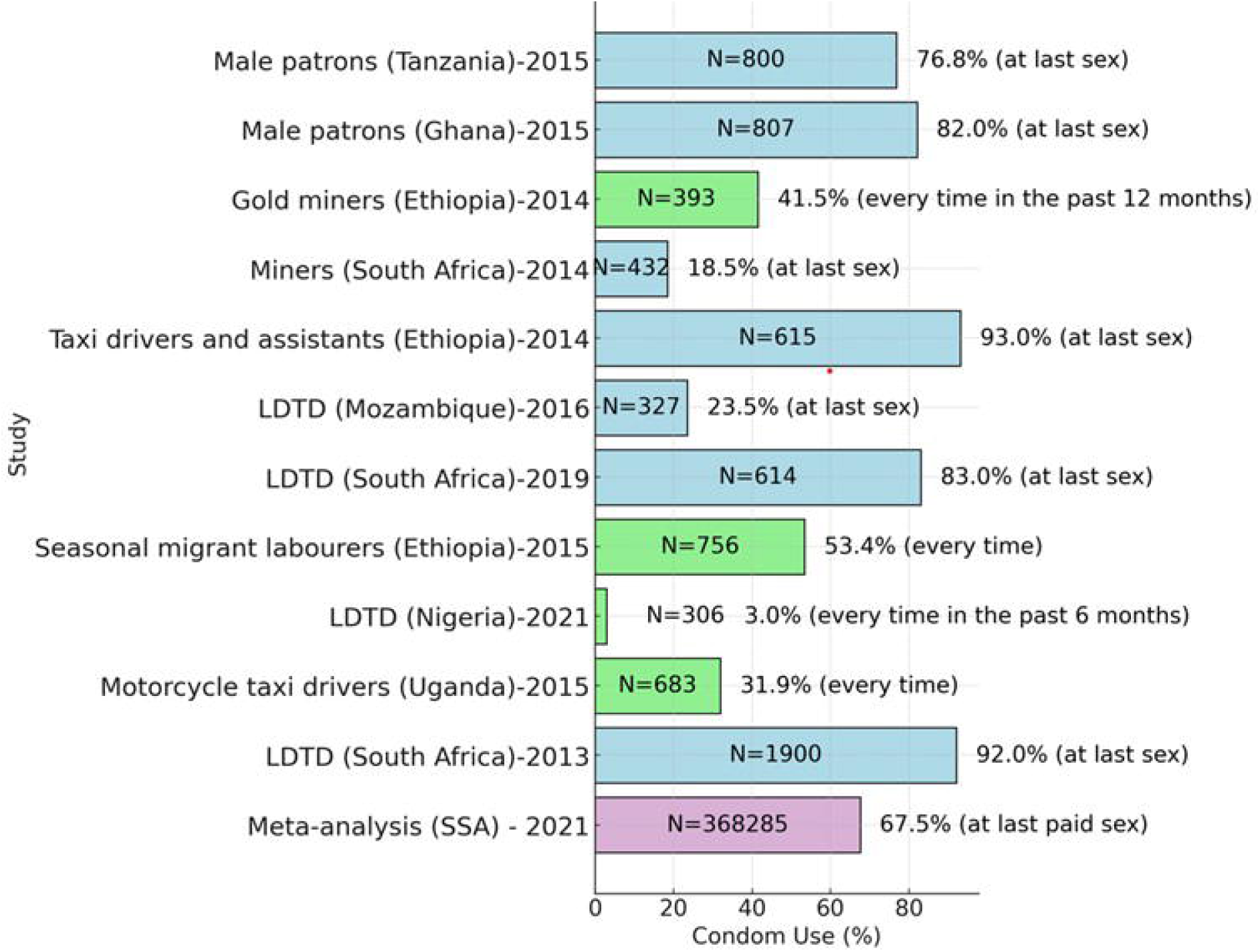
Condom use among men engaging with FSWs across studies in SSA

##### Findings from the high-risk subpopulation studies

Among men in high-risk sub-populations, reported condom use differed by sub-population and partner type. Condom use at the last sex ranged from 18.5% in 2014 to 93% in 2014(20, 31). Condom use in the past 6 months ranged from 22.6% in 2021 to 83.0 % 2019(17, 18). There was inconsistent condom use reported across these subgroups of men – Fig 4.

##### Factors associated with condom use

The meta-analysis did not report on factors associated with condom use among general population men. Limited condom use among LDTD was attributed to a lack of access to condoms. Although knowledge about HIV protection through condoms was reported to be high in some studies, this did not always translate to consistent use. Unsafe sex due to lack of access to condoms was also positively associated with higher HIV prevalence among these high-risk men.

#### HIV testing

##### Findings from the meta-analysis of the general population

The pooled proportion of men who ever tested for HIV among those who had ever had sex with FSWs was 64.9% (95% CI: 52.0%–75.9%) for surveys conducted between 2010 and 2020 – Fig 5. This increased from 31.9% (95% CI 25.5% - 39.1%) before 2010. No comparative data were reported for men who had never had sex with FSWs.

**Fig 5:**
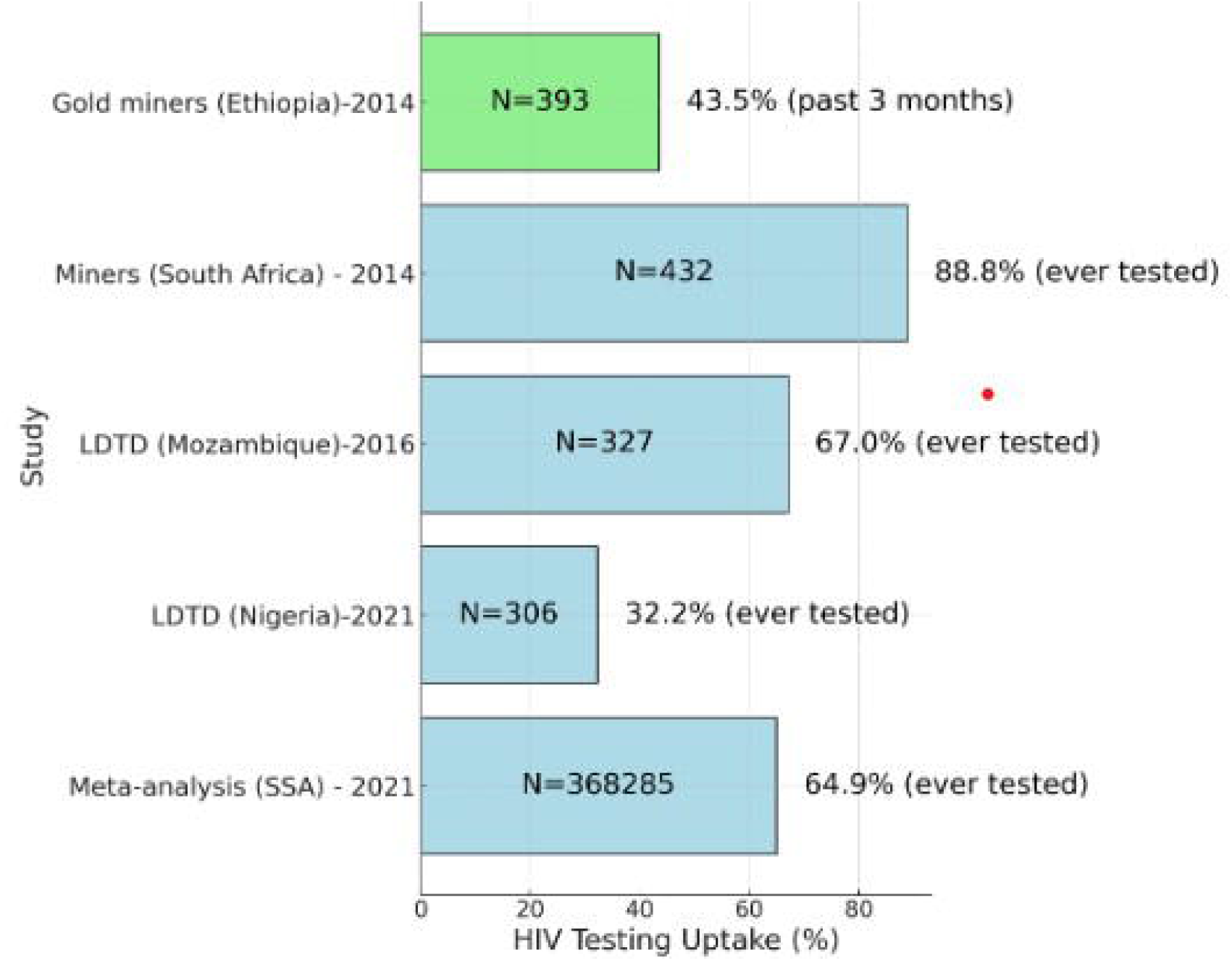
HIV testing uptake among men engaging with FSWs by study

##### Findings from the high-risk subpopulation studies

The proportion of subgroups of men who ever tested for HIV ranged from 36.9% in 2015 to 88.8% in 2014(19, 21, 31). Less than half, 43.5% of gold miners reported testing in the past 3 months in Ethiopia in 2014(23). A third of LDTD in Mozambique had never tested for HIV and thus lacked knowledge of their statuses in 2016(19) - Fig 5. These findings suggest that testing patterns were inconsistent across different countries and subgroups of high-risk men.

##### Factors associated with HIV testing

In the meta-analysis among general population men who reported ever sex with FSWs were more likely to have tested for HIV compared to men who did not. In the subgroups, limited use of health care services, access to health care services, and lack of educational interventions contributed to lower testing uptake rates. Fear and stigma also played a role in lower testing uptake. In the case of miners, higher uptake of testing resulted from a desire to know their status and testing as a requirement by the mine owners. LDTD in Mozambique who had never tested for HIV had significantly higher odds of being HIV positive compared to those who had ever tested for HIV, adjusted odds ratio [aOR: 2.2, 95% CI: 1.2 – 4.3)].

## Discussion

We found varied levels of paid sex between men in the general population and subgroups known to have higher HIV prevalence. Proportions of individuals reporting sex with an FSW varied among different subgroups. Factors associated with paid sex were not analyzed in the meta-analysis, though several factors were identified among the studies conducted in subgroups, but without comparative data for men who didn’t have sex with FSWs. Men who report having sex with FSWs have diverse socio-demographic and behavioral profiles. However, certain occupations pose higher risks for HIV and paid sex. HIV prevalence differed by population type. It was substantially higher among those who paid for sex compared to those who did not among men in the general population. HIV testing uptake was moderate in the general population but suboptimal in most subgroups.

Paying for sex per se was not identified as a risk factor for HIV among men from high-risk populations, suggesting either that paying for sex is ubiquitous (but underreported) or that a broader range of lifestyle factors contributes to a higher risk of HIV among these men.

One of the strengths of our study is the inclusion of a large meta-analysis conducted across 34 countries in SSA. The surveys were large, and representative, and the methods used were broadly similar making comparisons across regions meaningful. The geographical breadth provided insights into regional patterns in HIV epidemiology and sex with an FSW. Another strength of the study is the inclusion of studies conducted among different populations and geographies offering information on HIV prevalence among additional high-risk men.

Our review had limitations. One of the limitations of the review is that the meta-analysis did not provide a detailed analysis of the socio-demographic and behavioural characteristics associated with paid sex. In terms of the subpopulations we only identified a limited number of studies reporting research among high-risk men who have sex with FSWs reflecting the relatively small amount of research in this important group that has been conducted since 2010. We did not include unpublished or grey literature or articles that were not published in English (although we only found 1 article in other languages). Another drawback is that we were unable to capture the various social and cultural elements impacting sexual partnerships between FSWs and their sexual partners, which may have been recorded qualitatively, because the scoping review only included quantitative studies. The absence of disaggregated data on men who had sex with FSWs in comparison to those who did not is another drawback. Recall bias may have influenced the results, possibly resulting in missing, erroneous, or untrustworthy data that underreported or overreported sexual behaviour. Providing socially acceptable responses to sensitive information could also have affected study findings(34, 35). Subgroup studies employed a variety of methodological approaches for recruiting survey participants and used different measures of paying for sex across various settings. The lack of standardization in measuring some variables made it difficult to compare the included studies. One more challenge is the limited reporting of HIV prevalence data. This suggests that further research is needed to better understand the epidemiology of HIV among higher-risk men. This limited our ability to draw specific conclusions about FSW partners’ behaviors and needs and reduced the potential to tailor interventions specifically for this subgroup.

Although our findings are not generalizable, they can still inform local program development. There is no one-size-fits-all strategy that can effectively address the unique needs of these men.

Understanding HIV testing behaviors is crucial for men, as regular testing is key to early detection and treatment of HIV. Our findings indicate poor uptake of HIV testing among some men in the review. Comparably, 90.0% of migrant men in South Africa had ever tested for HIV in 2024 potentially due to increased awareness and access to testing services(36).

The lower uptake of HIV testing may be because of limited access to healthcare, fear of stigma and discrimination, limited knowledge about HIV risks, and economic challenges(37). Underreporting of engaging with sex workers by men may lead to a lower perceived risk of HIV, which is a barrier to testing(38). Embarrassment, fear of diagnosis, feeling healthy, fear of blood draws, distance from clinics, confidentiality concerns, and lack of testing knowledge could also contribute. Higher testing uptake could possibly be due to social desirability bias and desire to know their HIV statuses, which could have led to over-reporting of recent testing(31).

Future research should clarify definitions and variables of relevance, such as periods for having sex with an FSW, condom use, and accessing healthcare services, and separate findings between paying and non-paying sexual partnerships to better guide targeted HIV interventions. Furthermore, extensive multi-site studies using uniform protocols would enable significant cross-setting comparisons. Additionally, protocol standardization can enhance the creation of focused interventions by facilitating meta-analyses of HIV epidemiology among high-risk men, while longitudinal studies may aid in evaluating changes over time. It might be possible to update that review and include more recent surveys and more behavioural or other variables in the analysis enabling resource allocation to reduce the spread of HIV.

While the studies included in this review provide valuable insights into the behaviors and needs of men who have sex with FSWs and their engagement with HIV services, it is important to note that none of the included studies assessed the effectiveness of specific interventions. Therefore, the interventions discussed here are informed by observed risk behaviors and gaps in service use. However, further research is needed to evaluate their potential impact.

Targeted strategies are needed to address the social, economic, and behavioral factors driving these interactions. The International Labour Organization (ILO) emphasizes focusing on vulnerable places, such as land borders, trading posts, truck stops, and urban informal settlements, rather than vulnerable groups when tackling migration-related health concerns in the transport sector(39). Peer educators and trained community volunteers from high-risk groups can also promote healthier behaviors through behaviour change communication and condom distribution(40, 41).

HIV self-testing (HIVST) is crucial for populations facing stigma and discrimination, improving testing rates among hard-to-reach men(42). Innovative strategies, like digital technology and mobile testing, can increase HIV testing uptake. The use of chatbots and mobile apps increases information dissemination, reduces access barriers, and offers real-time support(43–47). Key opinion leaders also play important roles(48). Community and home-based interventions, with proven effectiveness, can increase testing among men who avoid healthcare facilities (49–53). Additionally, secondary distribution of self-test kits to women attending antenatal care and their partners can increase men’s testing rates(54, 55).

Programs using mobile clinics, such as Médecins Sans Frontières’ initiative in South Africa, increased ART initiation and testing among migrant farmworkers(56). Universal test and treat (UTT) and treatment as prevention (TasP) can further target high-risk men, including military personnel, with mandatory testing before deployments(57, 58). Multi-venue kit distribution in workplaces and social venues can offer convenience and privacy and successfully reach first-time testers(59, 60). Mobile testing at truck stops, military camps, and transport hubs further enhances HIV service access(61, 62).

Initiatives like North Star Alliance’s roadside clinics along transport corridors improve service access for truck drivers(63). The use of community ART distribution points (PODIs) and Community ART Refill Groups (CARGs) are effective for people living with HIV (PLWHIV) facing stigma by reducing waiting times and addressing accessibility issues for high-risk men(64, 65). Integrating PrEP and ART with sexual health services is crucial for these populations. Installing condom dispensers at public venues could help overcome barriers like stigma and privacy concerns, increasing uptake among men.

## Conclusions

Our study found varied prevalences of sex with FSWs among men, differing by population type. Furthermore, men who engage with FSWs cannot be easily identified by specific characteristics. Our results add critical information to the body of knowledge regarding men who are sexual partners of FSWs in SSA. In summary, the reviewed studies consistently showed that men who have sex with FSWs exhibited high-risk behaviors. Paying for sex was prevalent in studies involving men in occupations characterized by mobility and extended periods away from their spouses. Given the gaps in the literature on intervention effectiveness for these groups, future research should prioritize evaluating HIV prevention strategies tailored to men who have sex with FSWs. Finally, these findings highlight the importance of targeting subpopulations of men with interventions rather than solely relying on population-level data.

## Supporting information

PRISMA-ScR Checklist

## Supporting information

Additional Table 1: PRISMA-ScR Checklist

## Author Contributions

Conceptualization, G.M., J.H.; E.F. and F.C.; methodology, G.M.; P.M.; J.H.; E.F.; and F.C.; results, G.M.; discussion, G.M.; J.H.; E.F.; and F.C.; conclusion, G.M.; writing—original draft preparation, G.M.; writing—review and editing, G.M., J.H.; E.F. and F.C. All authors have read and agreed to the published version of the manuscript.

## Institutional Review Board Statement

Not applicable.

## Informed Consent Statement

Not applicable.

## Data Availability Statement

Since this study is a scoping review, there is no statistical data set.

## Public Involvement Statement

No public involvement in any aspect of this research.

## Guidelines and Standards Statement

This manuscript was drafted against the Preferred Reporting Items of Systematic Reviews and Meta-analyses guidelines for review research.

## S1 APPENDIX: Appraisal tool for Cross-sectional Studies (AXIS)

|  |  |  |  |  |
| --- | --- | --- | --- | --- |
| **1. Alcohol use and sexual risk behaviours in the Armed Forces of the Democratic Republic of Congo (DRC)** |  |  |  |  |
|  | **Question** | **Yes** | **No** | **Don't know/Comment** |
| ***Introduction*** |  |  |  |  |
| 1 | Were the aims/objectives of the study clear? | Yes |  |  |
| ***Methods*** |  |  |  |  |
| 2 | Was the study design appropriate for the stated aim(s)? |  | No | Convenience sample is non-probabilistic |
| 3 | Was the sample size justified? |  | No |  |
| 4 | Was the target/reference population clearly defined? (Is it clear who the research was about?) | Yes |  |  |
| 5 | Was the sample frame taken from an appropriate population base so that it closely represented the target/reference population under investigation? |  | No | Non-representative because of biased selection |
| 6 | Was the selection process likely to select subjects/participants that were representative of the target/reference population under investigation? |  | No |  |
| 7 | Were measures undertaken to address and categorise non-responders? |  | No |  |
| 8 | Were the risk factor and outcome variables measured appropriate to the aims of the study? | Yes |  |  |
| 9 | Were the risk factor and outcome variables measured correctly using instruments/measurements that had been trialled, piloted or published previously? |  |  | I do not know since its not stated |
| 10 | Is it clear what was used to determined statistical significance and/or precision estimates? (e.g. p-values, confidence intervals) | Yes |  |  |
| 11 | Were the methods (including statistical methods) sufficiently described to enable them to be repeated? | Yes |  |  |
| **Results** |  |  |  |  |
| 12 | Were the basic data adequately described? | Yes |  |  |
| 13 | Does the response rate raise concerns about non-response bias? |  | No |  |
| 14 | If appropriate, was information about non-responders described? |  | No |  |
| 15 | Were the results internally consistent? | Yes |  |  |
| 16 | Were the results presented for all the analyses described in the methods? | Yes |  |  |
| **Discussion** |  |  |  |  |
| 17 | Were the authors' discussions and conclusions justified by the results? | Yes |  |  |
| 18 | Were the limitations of the study discussed? | Yes |  |  |

| **Other** |  |  |  |  |
| --- | --- | --- | --- | --- |
| 19 | Were there any funding sources or conflicts of interest that may affect the authors' interpretation of the results? |  | No | The authors declared no conflicts of interest |
| 20 | Was ethical approval or consent of participants attained? | Yes |  |  |

| **2. High-risk motorcycle taxi drivers in the HIV/AIDS era: a respondent-driven sampling survey in Kampala, Uganda** |  |  |  |  |
| --- | --- | --- | --- | --- |
|  | **Question** | **Yes** | **No** | **Don't know/ Comment** |
| **Introduction** |  |  |  |  |
| 1 | Were the aims/objectives of the study clear? | Yes |  |  |
| **Methods** |  |  |  |  |
| 2 | Was the study design appropriate for the stated aim(s)? | Yes | o | Network population |
| 3 | Was the sample size justified? | Yes |  | Sample size calculation conducted |
| 4 | Was the target/reference population clearly defined? (Is it clear who the research was about?) | Yes |  |  |
| 5 | Was the sample frame taken from an appropriate population base so that it closely represented the target/reference population under investigation? | Yes |  |  |
|  | Was the selection process likely to select | Yes |  |  |
| 6 | subjects/participants that were representative of the target/reference population under investigation? |  |  |  |
| 7 | Were measures undertaken to address and categorise non-responders? |  | No |  |
| 8 | Were the risk factor and outcome variables measured appropriate to the aims of the study? | Yes |  |  |
| 9 | Were the risk factor and outcome variables measured correctly using instruments/measurements that had been trialled, piloted, or published previously? | Yes |  |  |
| 10 | Is it clear what was used to determine statistical significance and/or precision estimates? (e.g. p-values, confidence intervals) | Yes |  |  |
| 11 | Were the methods (including statistical methods) sufficiently described to enable them to be repeated? | Yes |  |  |
| **Results** |  |  |  |  |
| 12 | Were the basic data adequately described? | Yes |  |  |
| 13 | Does the response rate raise concerns about non-response bias? |  | No |  |
| 14 | If appropriate, was information about non-responders described? |  | No |  |
| 15 | Were the results internally consistent? | Yes |  |  |
| 16 | Were the results presented for all the analyses described in the methods? | Yes |  |  |

| ***Discussion*** |  |  |  |  |
| --- | --- | --- | --- | --- |
| 17 | Were the authors' discussions and conclusions justified by the results? | Yes |  |  |
| 18 | Were the limitations of the study discussed? | Yes |  |  |
| ***Other*** |  |  |  |  |
| 19 | Were there any funding sources or conflicts of interest that may affect the authors' interpretation of the results? |  | No | The authors declared no conflicts of interest |
| 20 | Was ethical approval or consent of participants attained? | Yes |  |  |

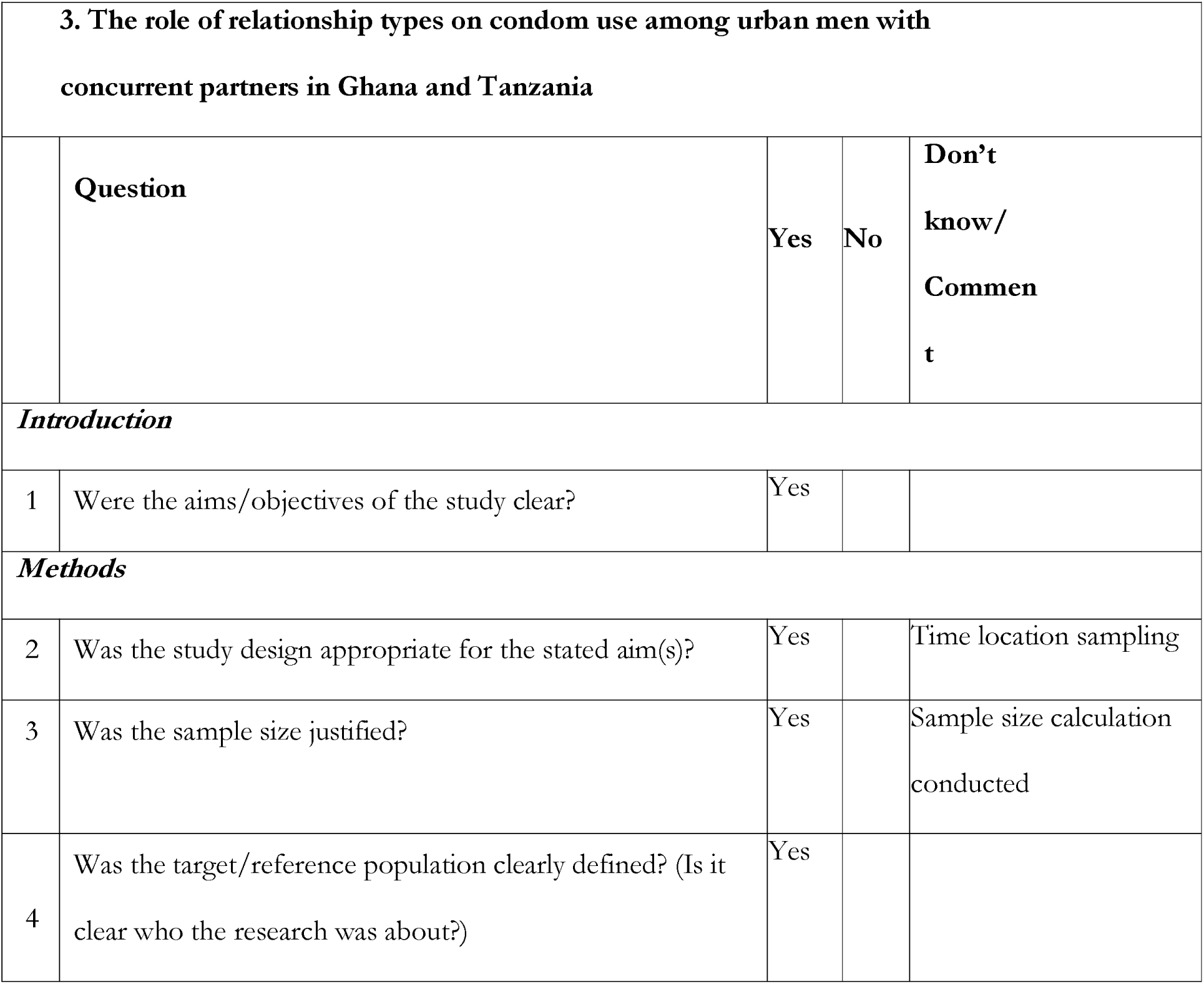
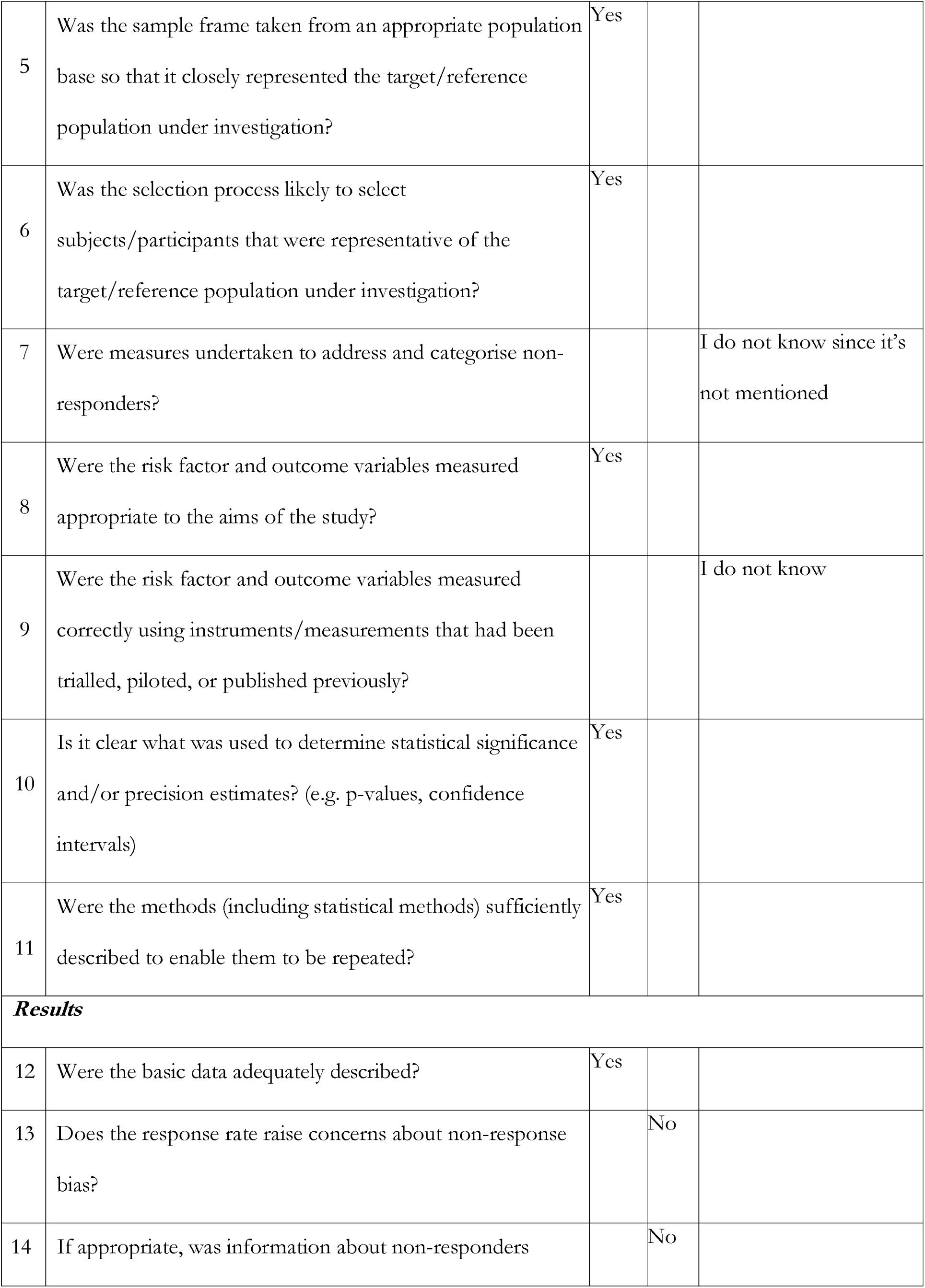
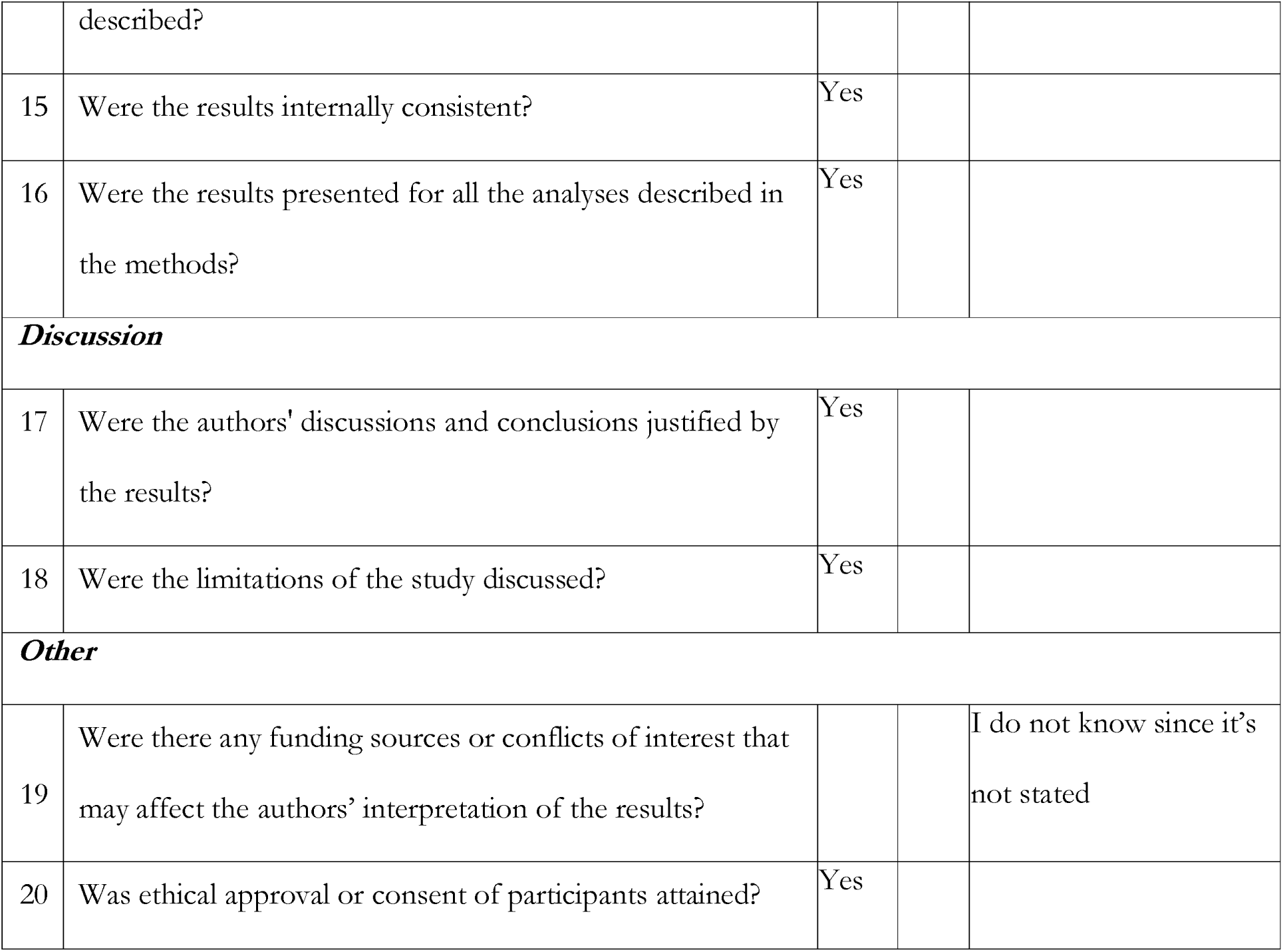

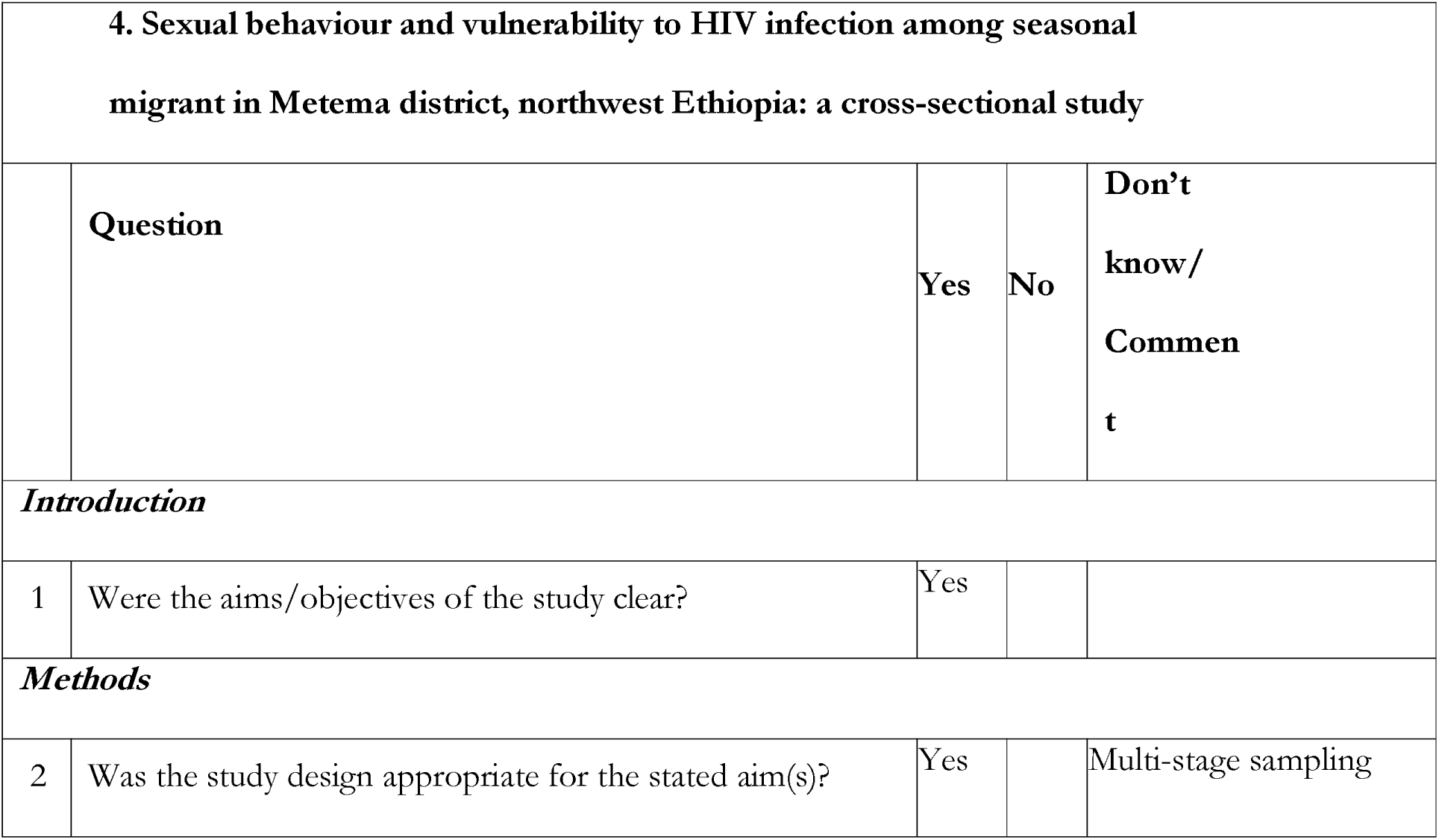
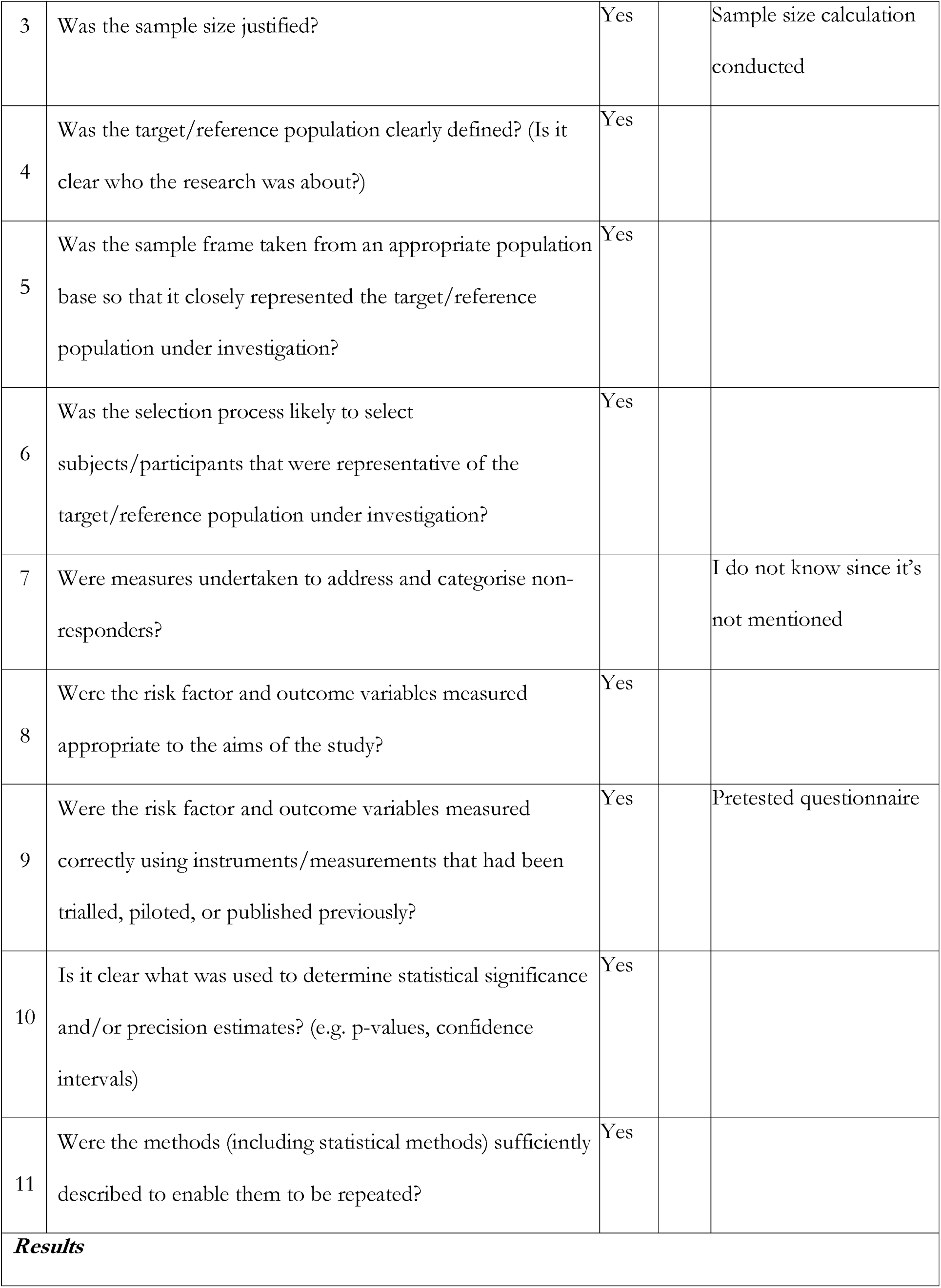
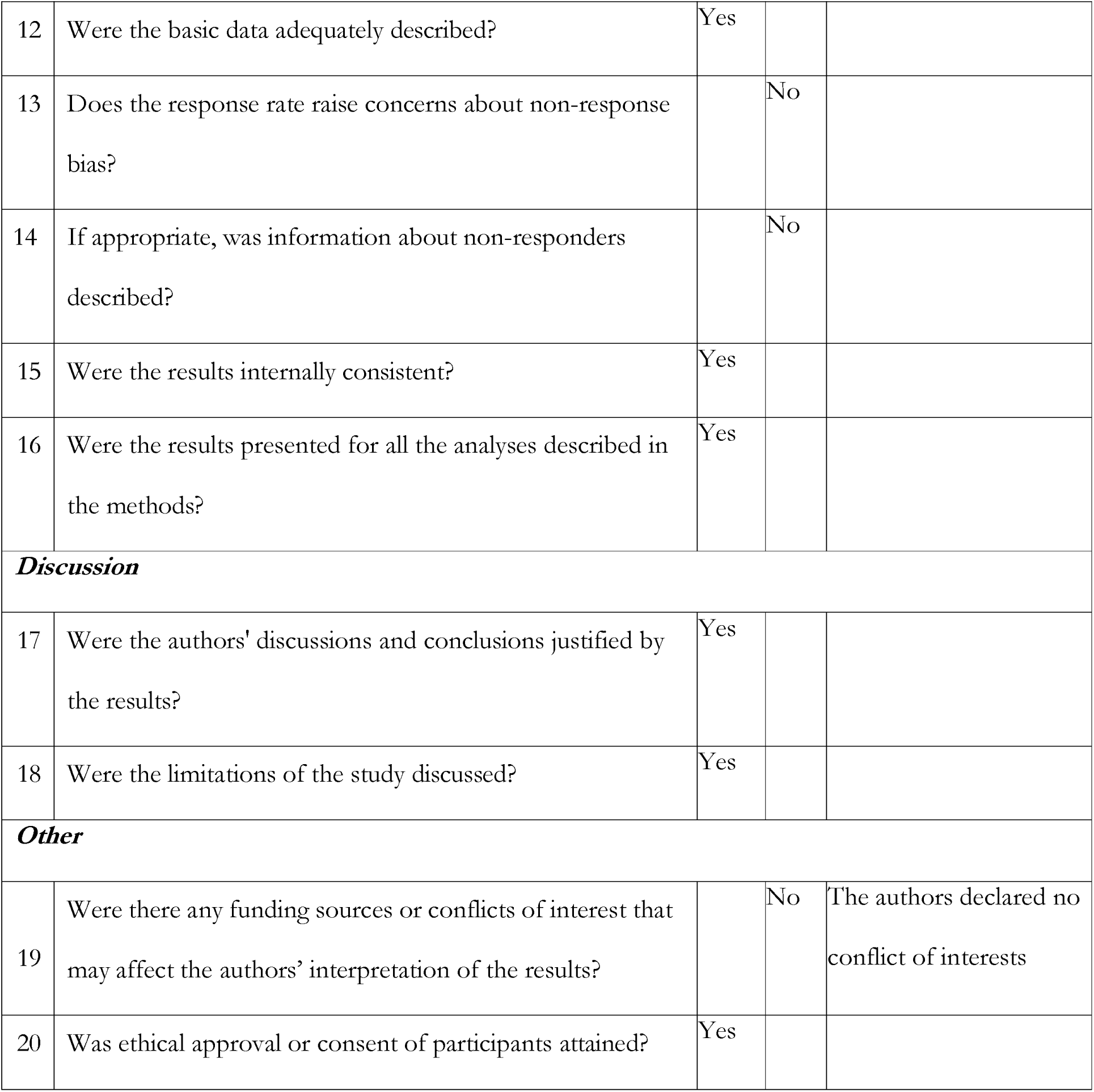

| **5. HIV related risk behaviours among taxi drivers and their assistants in Addis Ababa, Ethiopia: descriptive cross-sectional survey** |  |  |  |  |
| --- | --- | --- | --- | --- |
|  | **Question** | **Yes** | **No** | **Don't know/ Comment** |
| ***Introduction*** |  |  |  |  |
| 1 | Were the aims/objectives of the study clear? | Yes |  |  |
| ***Methods*** |  |  |  |  |
| 2 | Was the study design appropriate for the stated aim(s)? | Yes |  | Multi-stage sampling |
| 3 | Was the sample size justified? | Yes |  | Sample size calculation conducted |
| 4 | Was the target/reference population clearly defined? (Is it clear who the research was about?) | Yes |  |  |
| 5 | Was the sample frame taken from an appropriate population base so that it closely represented the target/reference population under investigation? | Yes |  |  |
| 6 | Was the selection process likely to select subjects/participants that were representative of the target/reference population under investigation? | Yes |  |  |
| 7 | Were measures undertaken to address and categorise non-responders? |  |  | I do not know since it's not mentioned |
| 8 | Were the risk factor and outcome variables measured appropriate to the aims of the study? | Yes |  |  |
| 9 | Were the risk factor and outcome variables measured correctly using instruments/measurements that had been trialled, piloted, or published previously? | Yes |  | Pretested questionnaire |
| 10 | Is it clear what was used to determine statistical significance and/or precision estimates? (e.g. p-values, confidence intervals) | Yes |  |  |
| 11 | Were the methods (including statistical methods) sufficiently described to enable them to be repeated? | Yes |  |  |
| **Results** |  |  |  |  |
| 12 | Were the basic data adequately described? | Yes |  |  |
| 13 | Does the response rate raise concerns about non-response bias? |  | No |  |
| 14 | If appropriate, was information about non-responders described? |  | No |  |
| 15 | Were the results internally consistent? | Yes |  |  |
| 16 | Were the results presented for all the analyses described in the methods? | Yes |  |  |
| **Discussion** |  |  |  |  |
| 17 | Were the authors' discussions and conclusions justified by the results? | Yes |  |  |
| 18 | Were the limitations of the study discussed? | Yes |  |  |
| **Other** |  |  |  |  |
| 19 | Were there any funding sources or conflicts of interest that may affect the authors' interpretation of the results? |  | No | The authors declared no conflict of interests |
| 20 | Was ethical approval or consent of participants attained? | Yes |  |  |

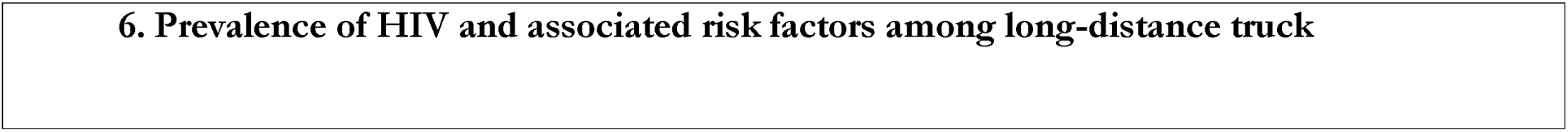
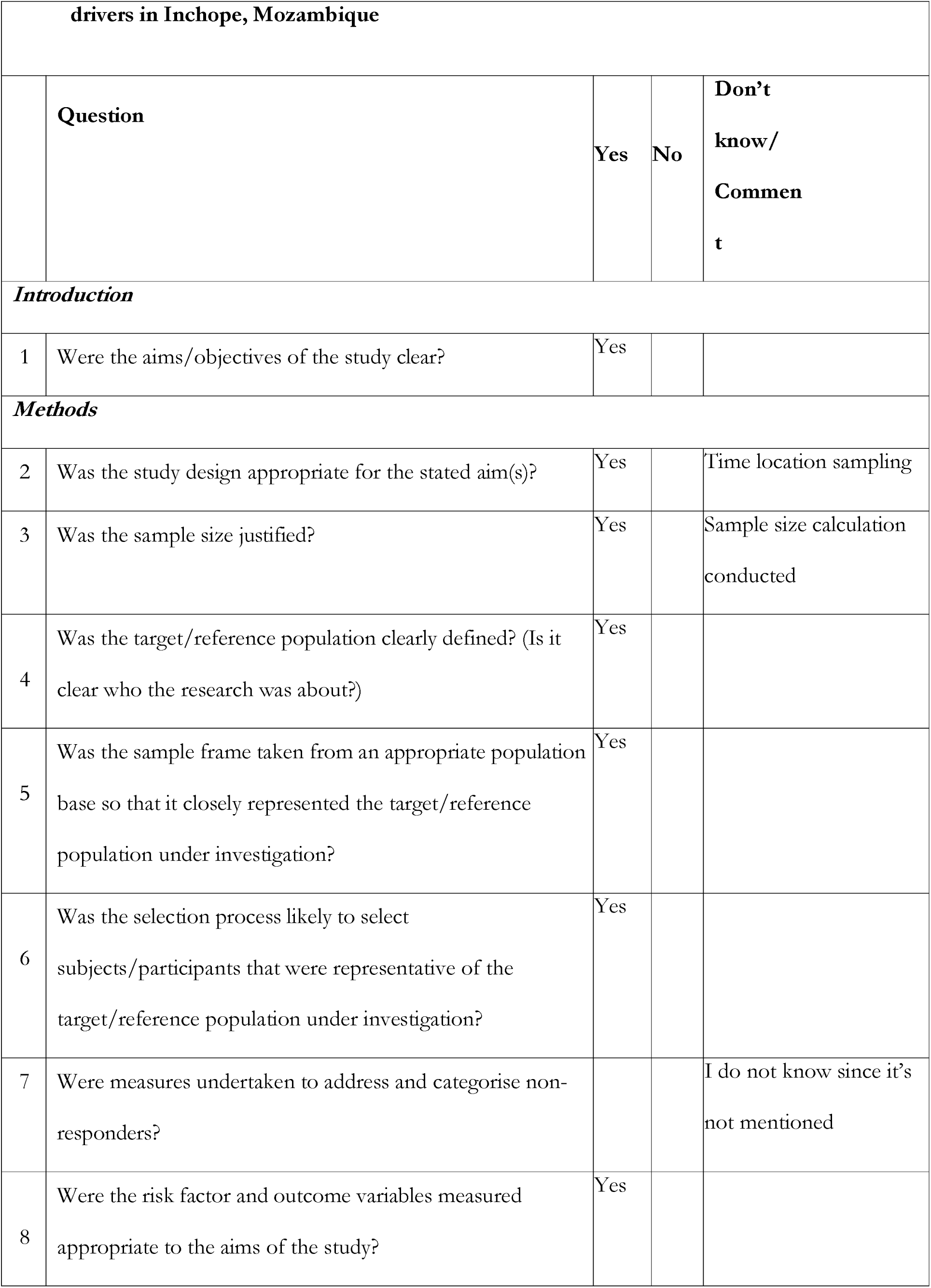
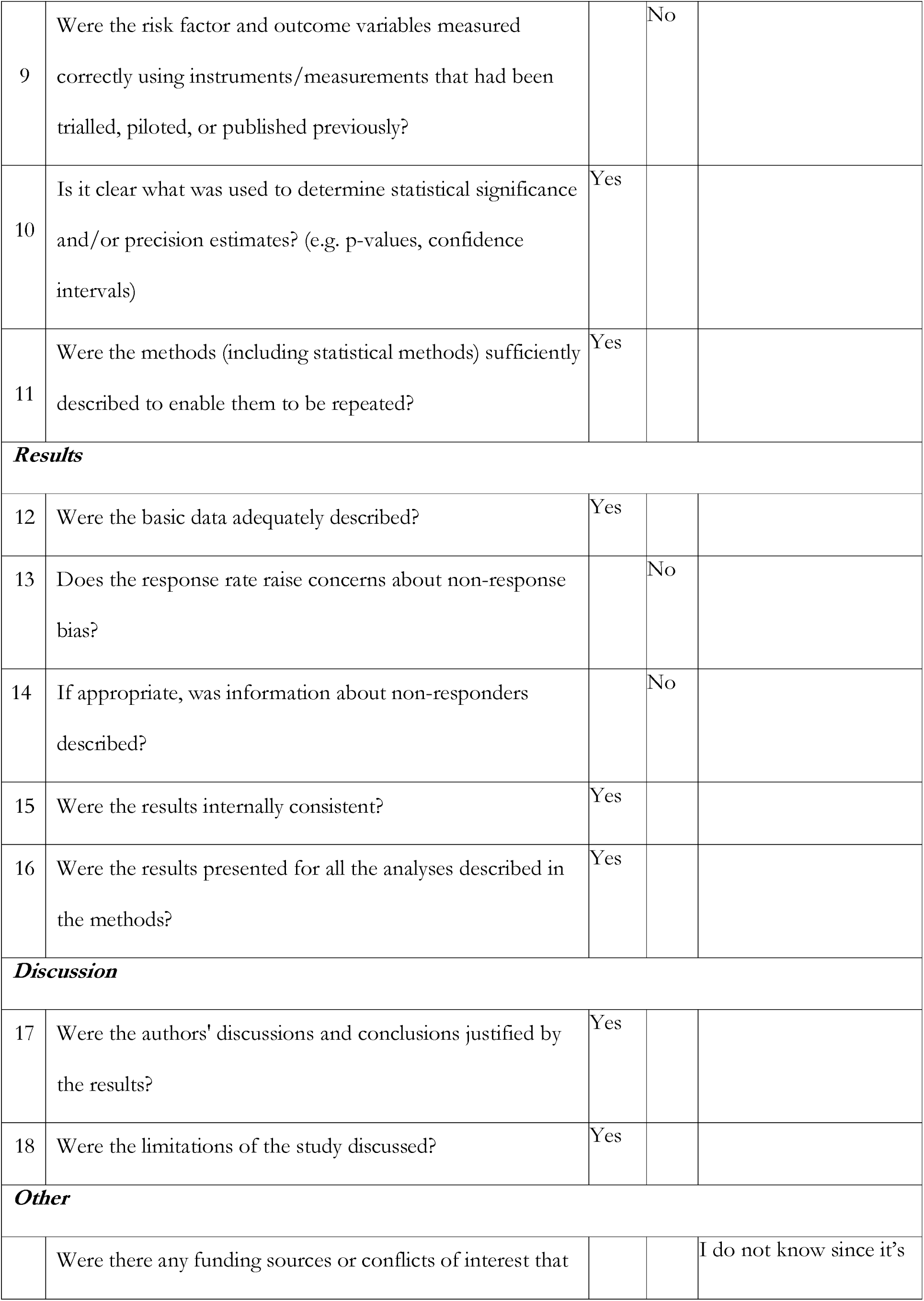
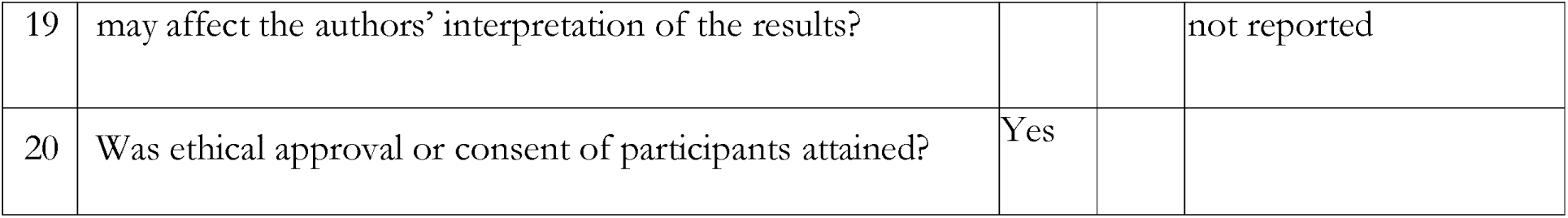

6. Prevalence of HIV and associated risk factors among long-distance truck
| **drivers in Inchope, Mozambique** |  |  |  |  |
| --- | --- | --- | --- | --- |
|  | **Question** | **Yes** | **No** | **Don't know/**<br>**Comment** |
| ***Introduction*** |  |  |  |  |
| 1 | Were the aims/objectives of the study clear? | Yes |  |  |
| ***Methods*** |  |  |  |  |
| 2 | Was the study design appropriate for the stated aim(s)? | Yes |  | Time location sampling |
| 3 | Was the sample size justified? | Yes |  | Sample size calculation conducted |
| 4 | Was the target/reference population clearly defined? (Is it clear who the research was about?) | Yes |  |  |
| 5 | Was the sample frame taken from an appropriate population base so that it closely represented the target/reference population under investigation? | Yes |  |  |
| 6 | Was the selection process likely to select subjects/participants that were representative of the target/reference population under investigation? | Yes |  |  |
| 7 | Were measures undertaken to address and categorise non-responders? |  |  | I do not know since it's not mentioned |
| 8 | Were the risk factor and outcome variables measured appropriate to the aims of the study? | Yes |  |  |
| 9 | Were the risk factor and outcome variables measured correctly using instruments/measurements that had been trialled, piloted, or published previously? |  | No |  |
| 10 | Is it clear what was used to determine statistical significance and/or precision estimates? (e.g. p-values, confidence intervals) | Yes |  |  |
| 11 | Were the methods (including statistical methods) sufficiently described to enable them to be repeated? | Yes |  |  |
| **Results** |  |  |  |  |
| 12 | Were the basic data adequately described? | Yes |  |  |
| 13 | Does the response rate raise concerns about non-response bias? |  | No |  |
| 14 | If appropriate, was information about non-responders described? |  | No |  |
| 15 | Were the results internally consistent? | Yes |  |  |
| 16 | Were the results presented for all the analyses described in the methods? | Yes |  |  |
| **Discussion** |  |  |  |  |
| 17 | Were the authors' discussions and conclusions justified by the results? | Yes |  |  |
| 18 | Were the limitations of the study discussed? | Yes |  |  |
| **Other** |  |  |  |  |
|  | Were there any funding sources or conflicts of interest that |  |  | I do not know since it's |
| 19 | may affect the authors' interpretation of the results? |  |  | not reported |
| 20 | Was ethical approval or consent of participants attained? | Yes |  |  |

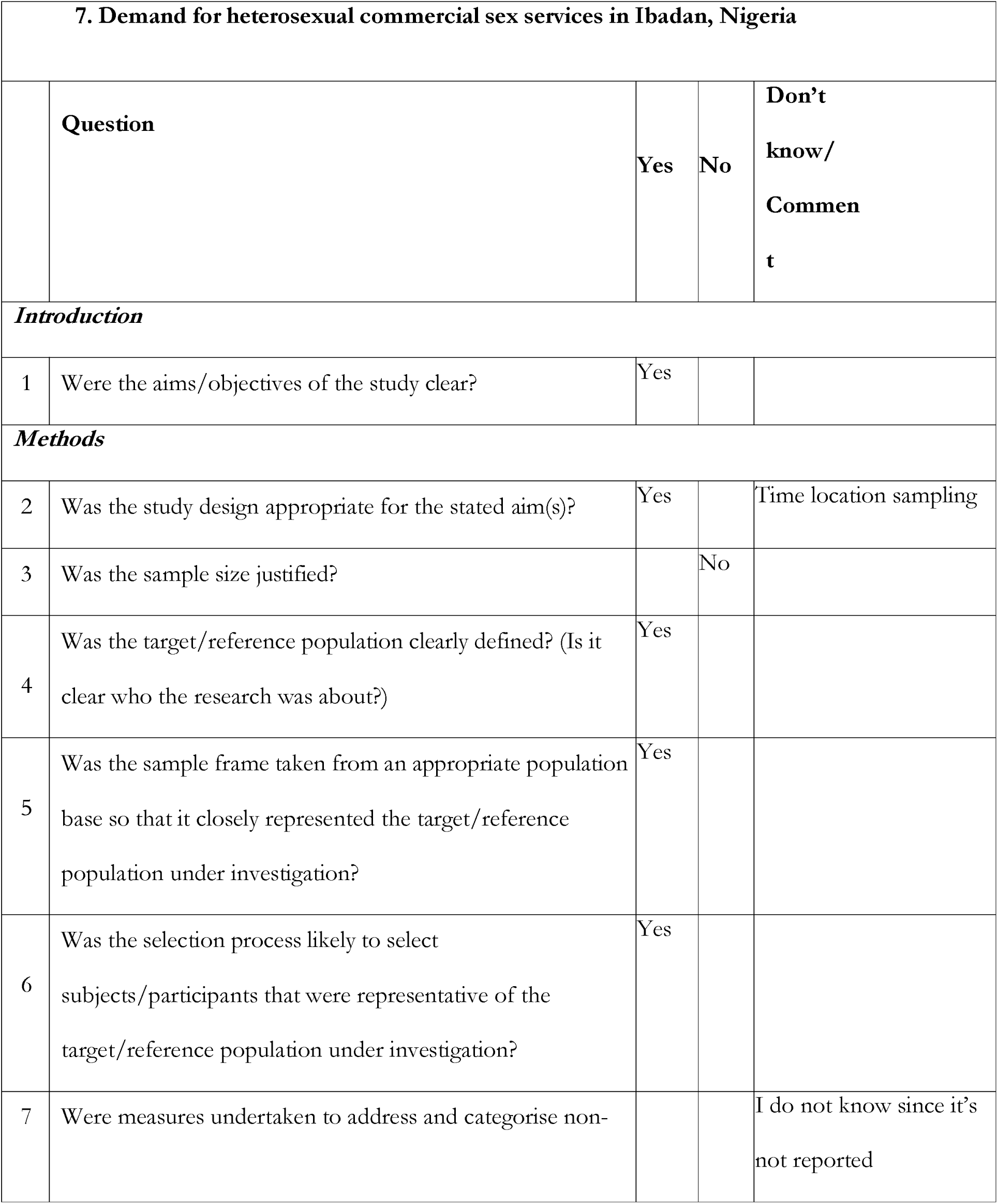
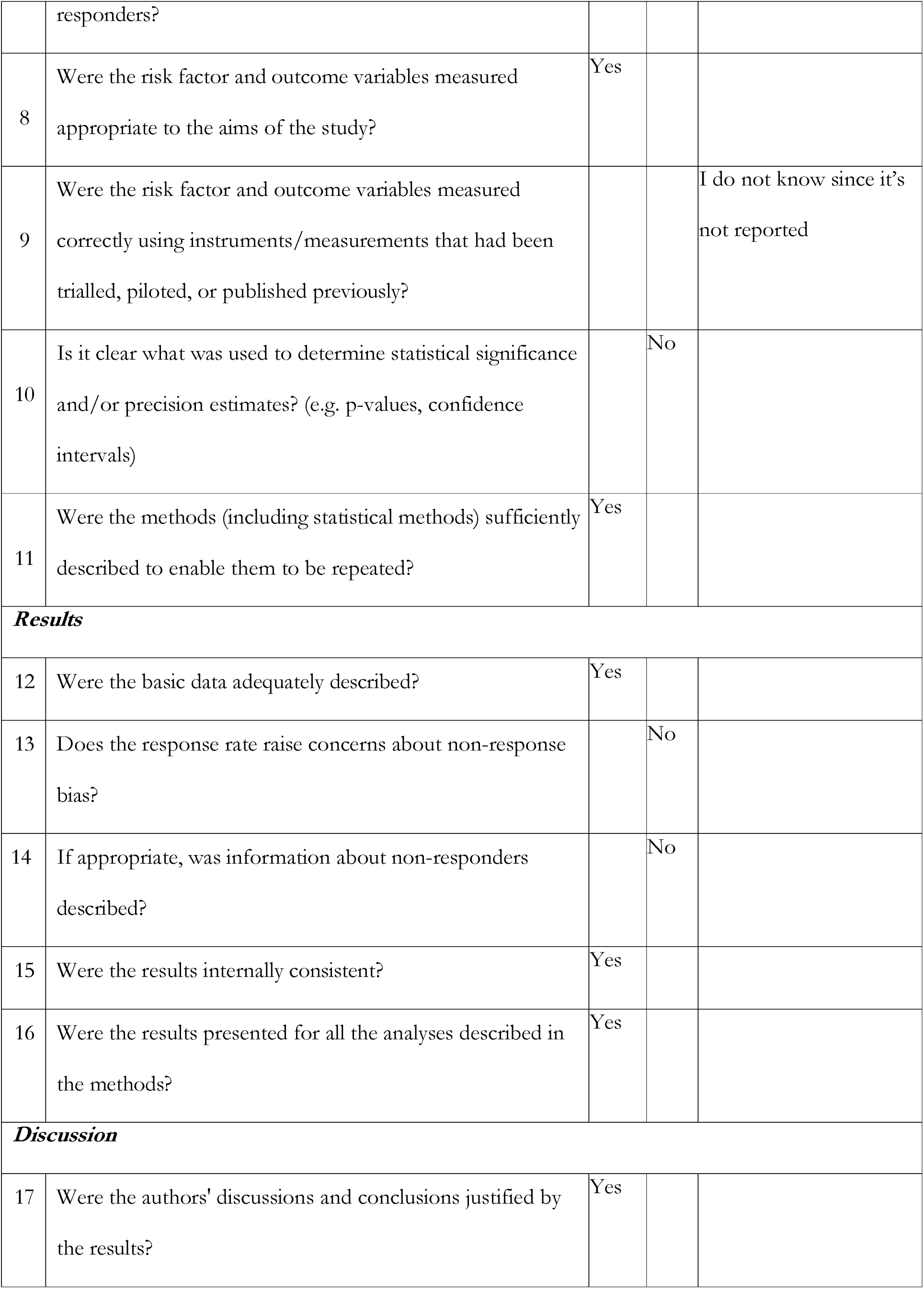
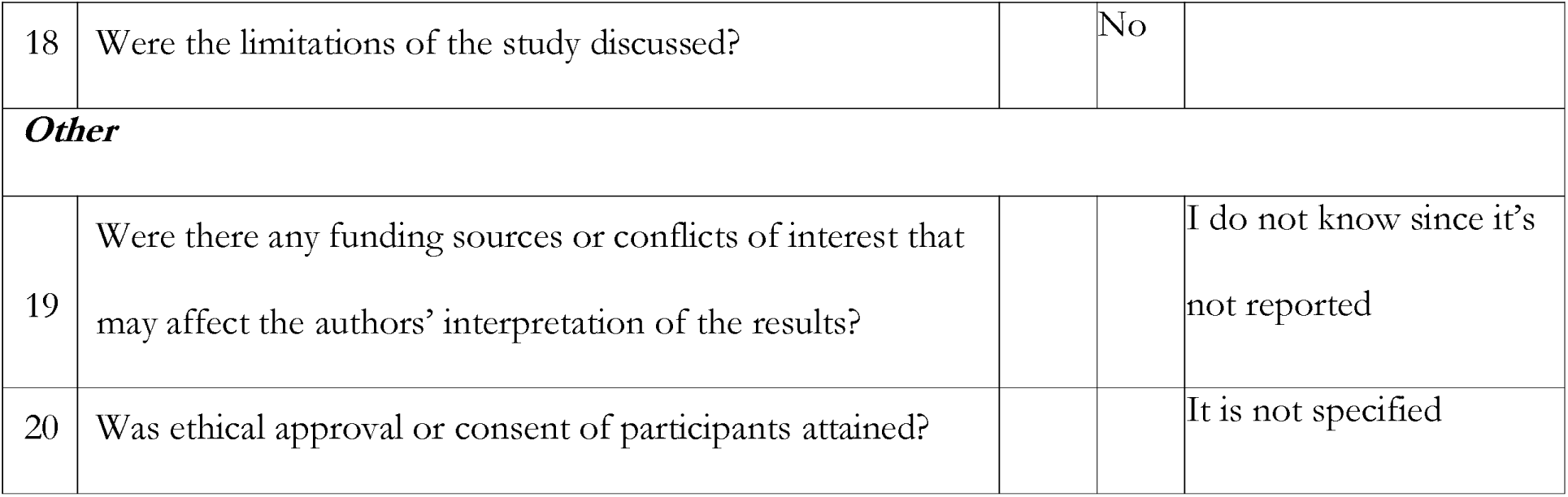

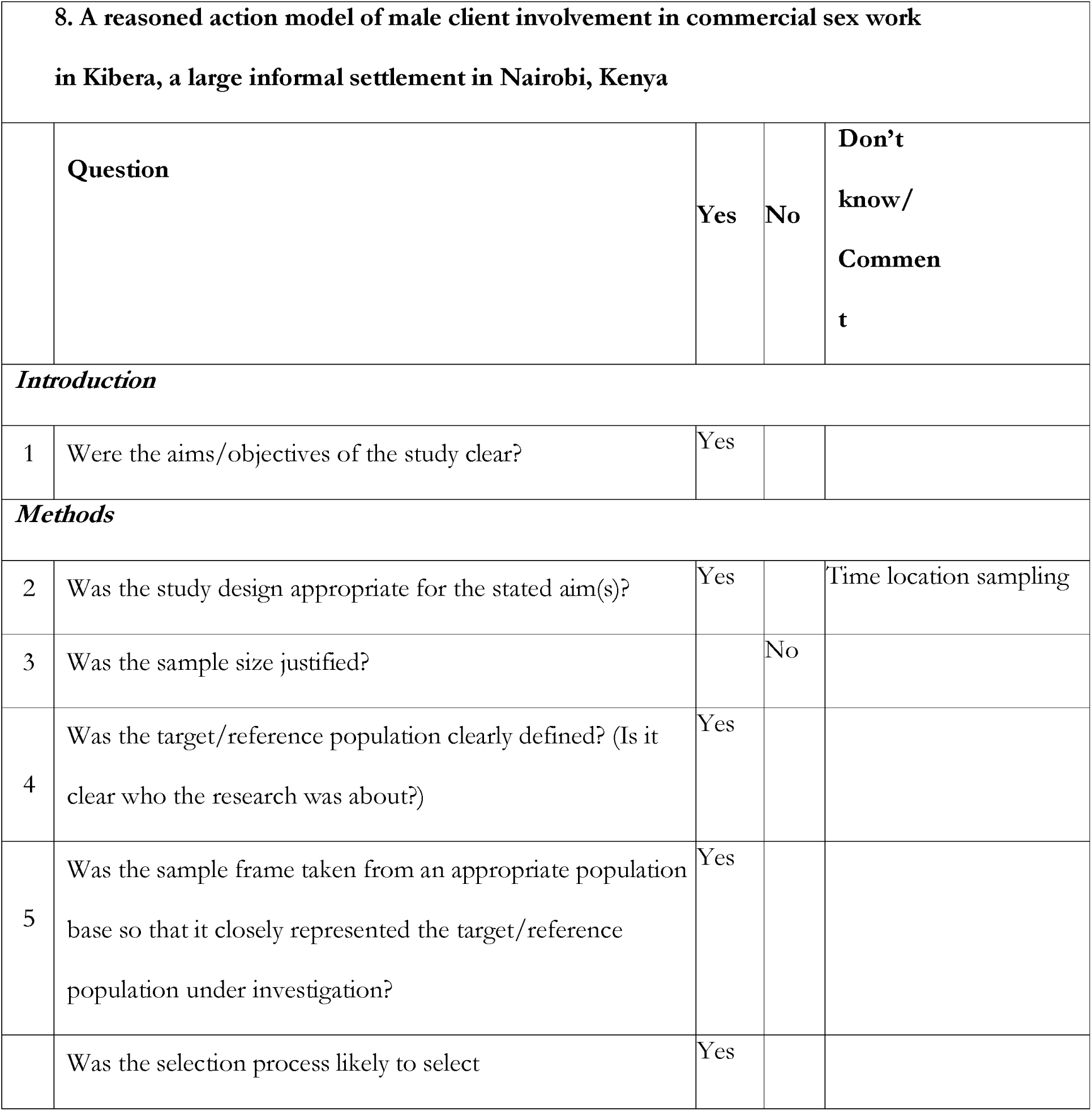
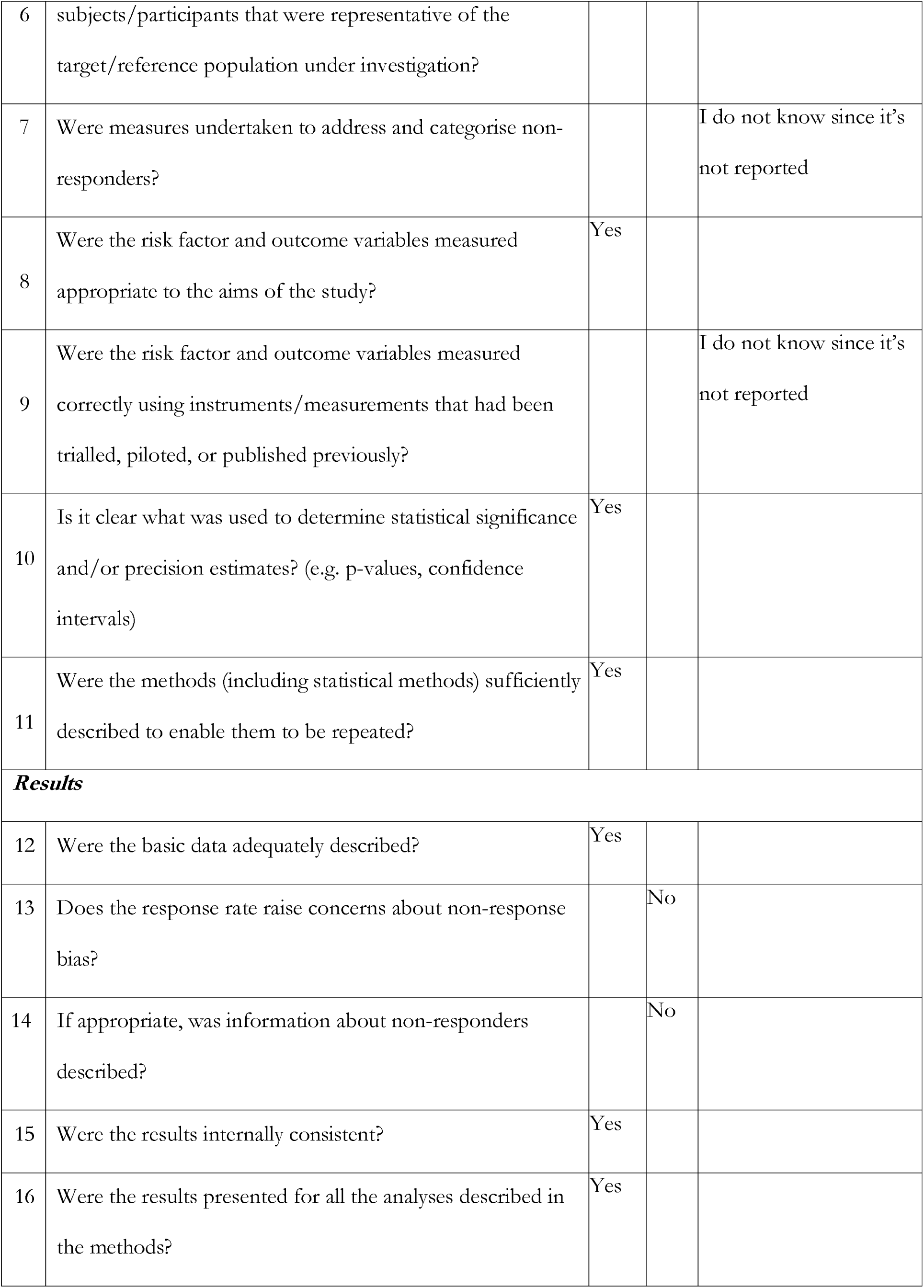
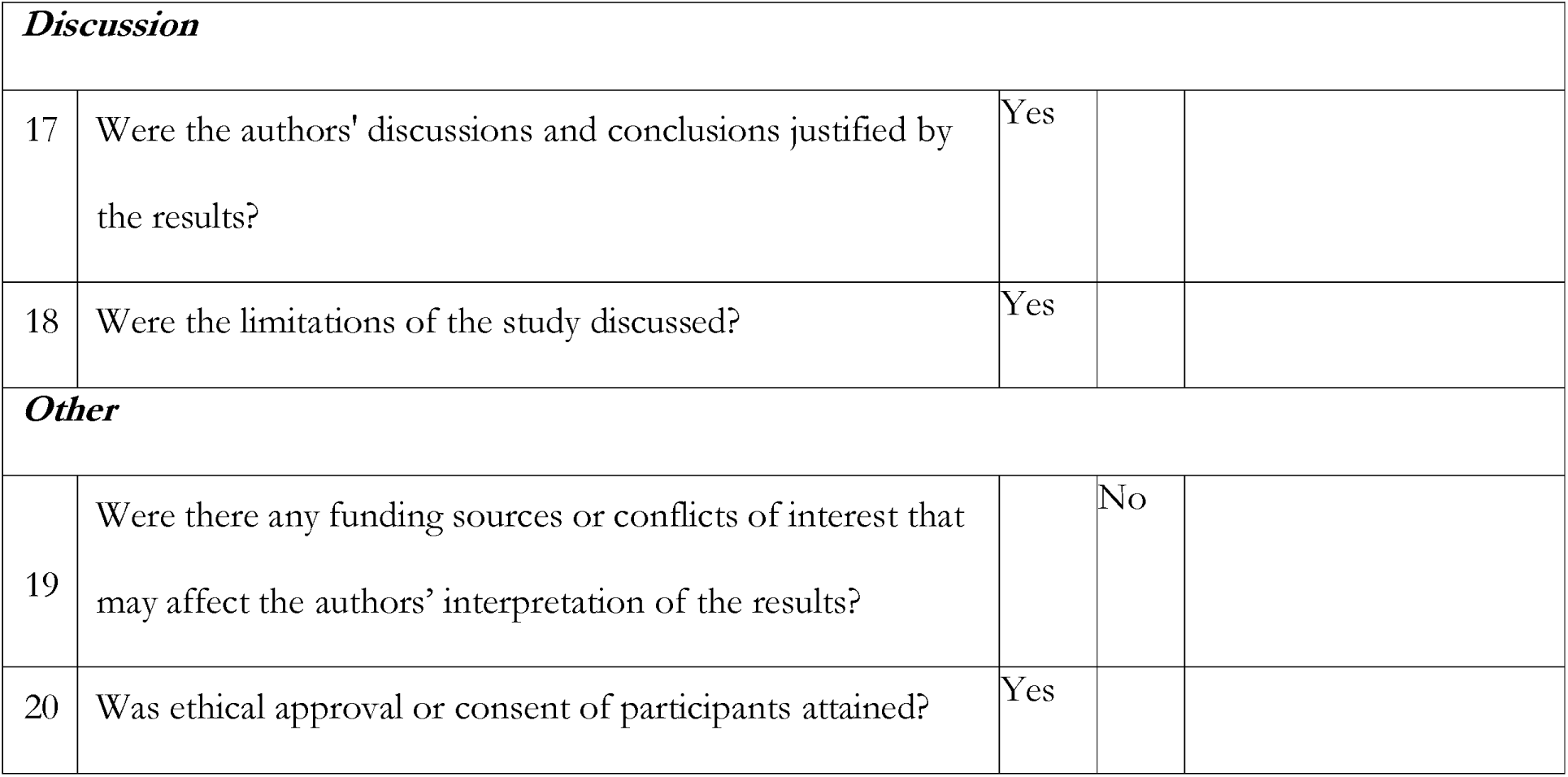

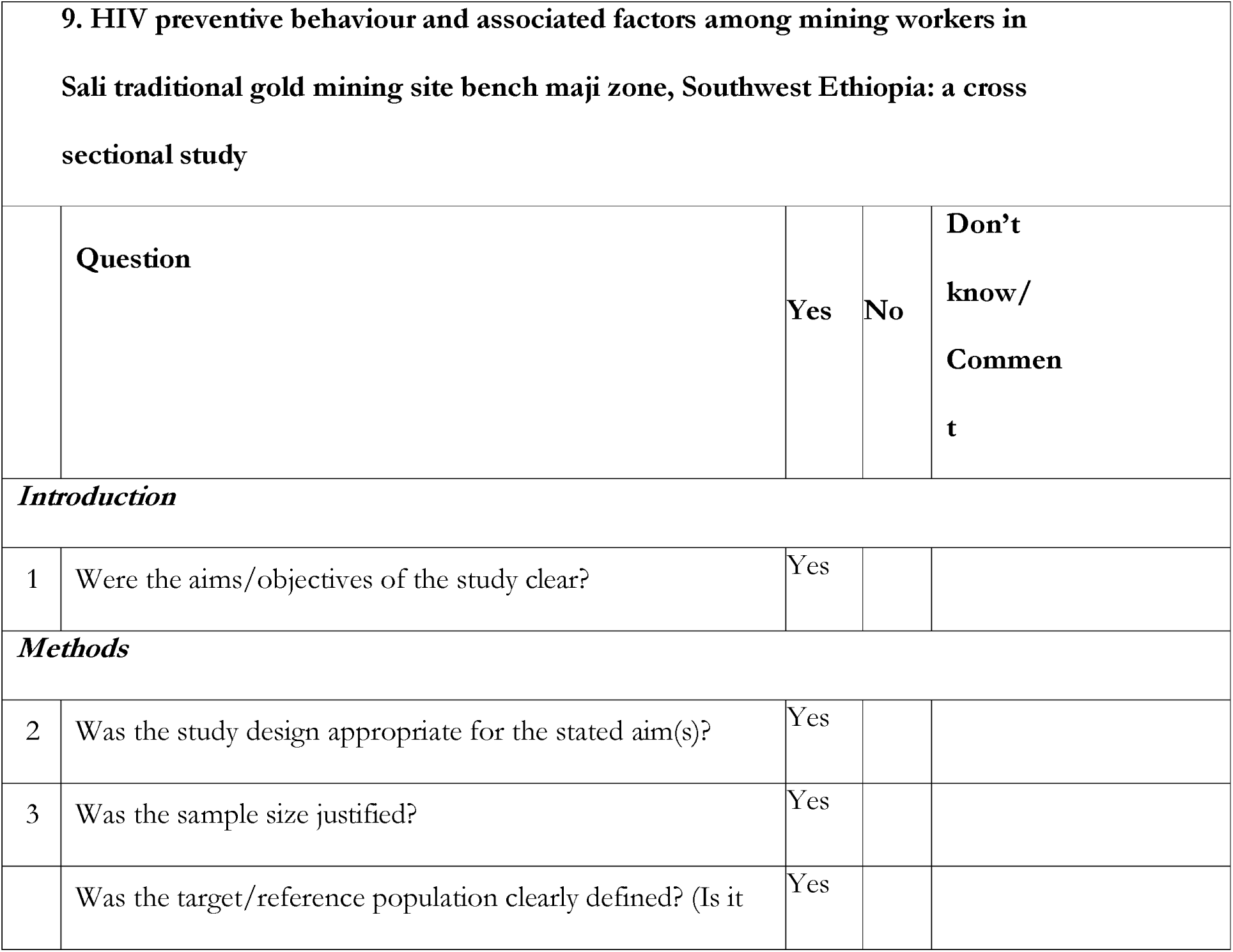
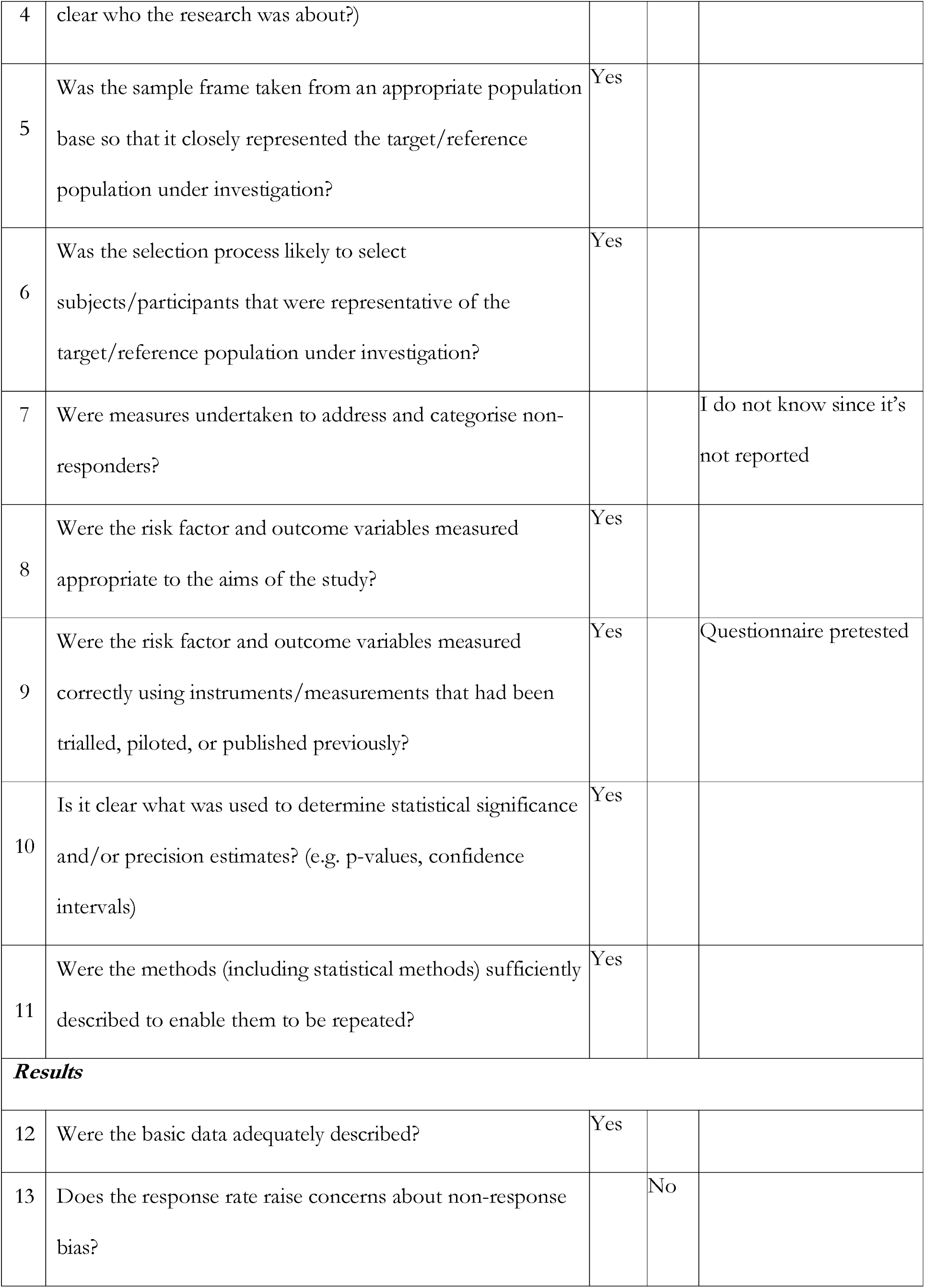
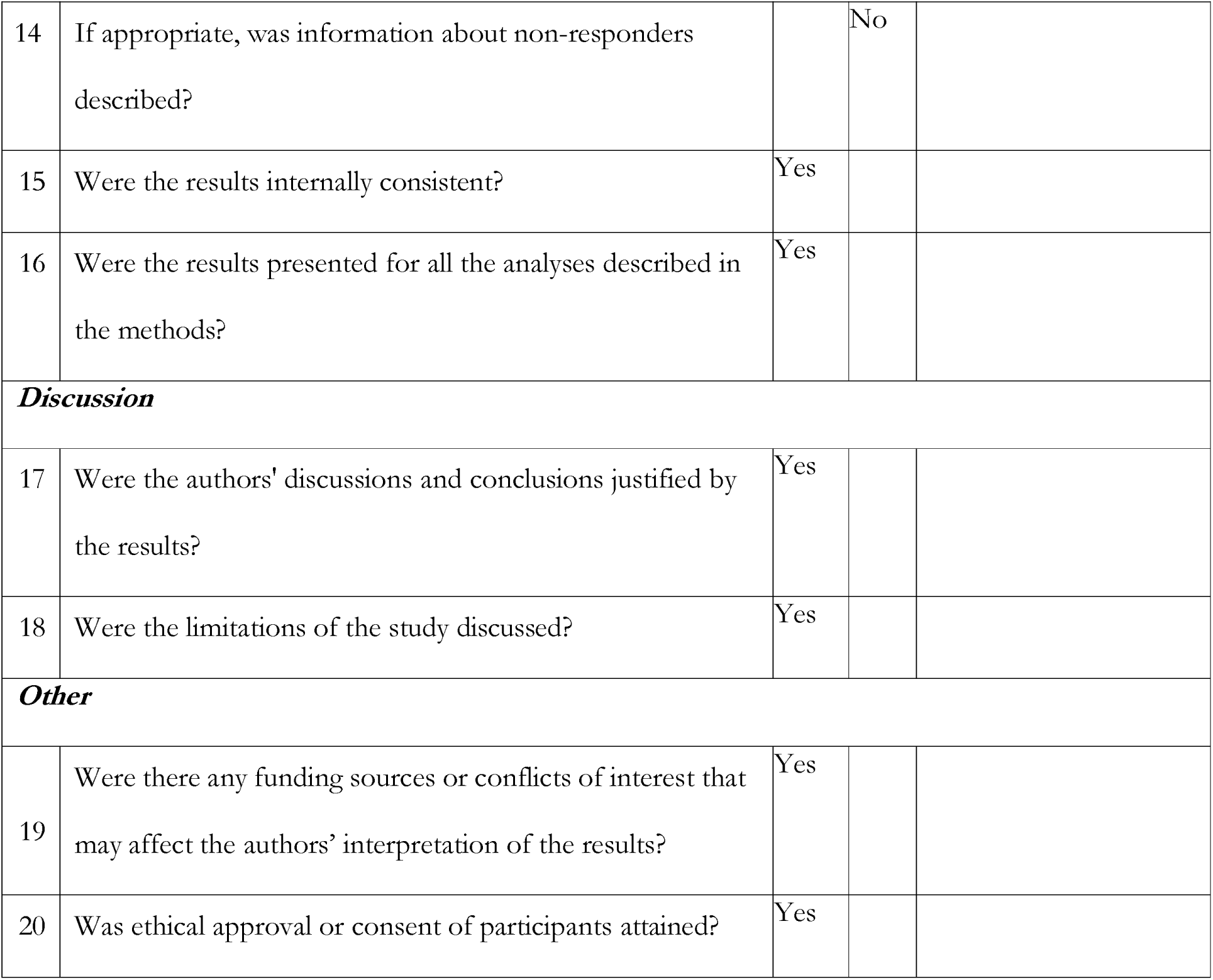

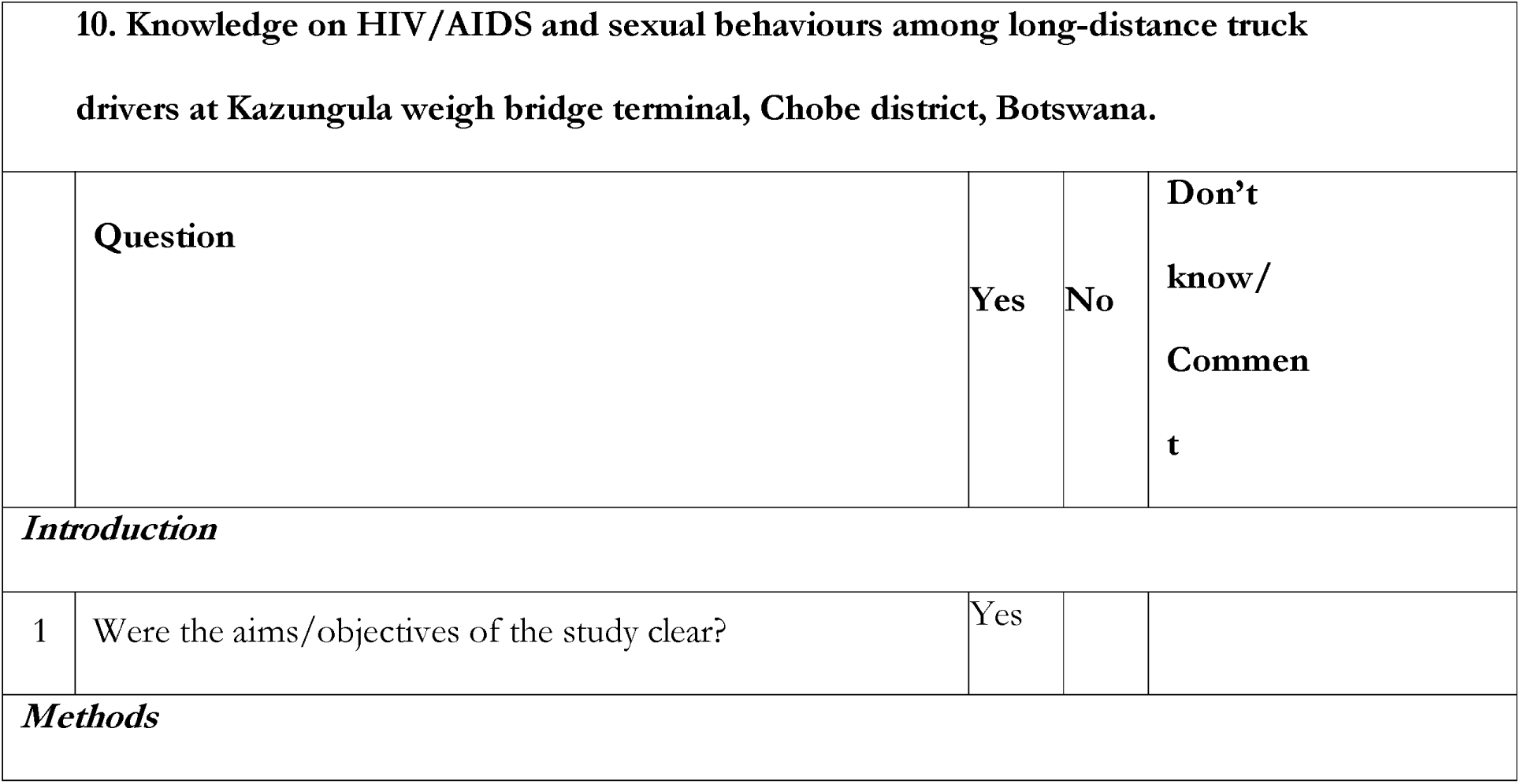
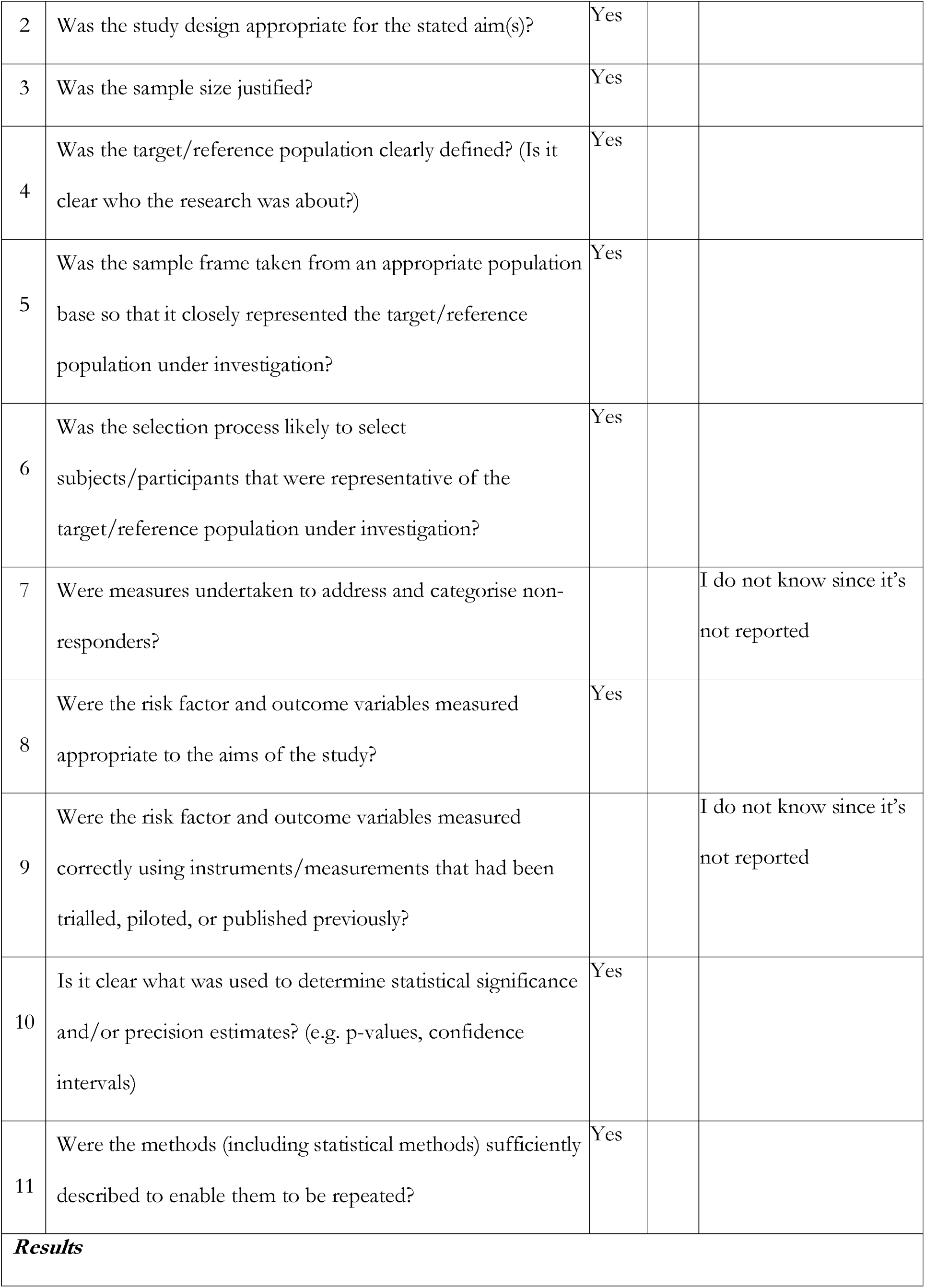
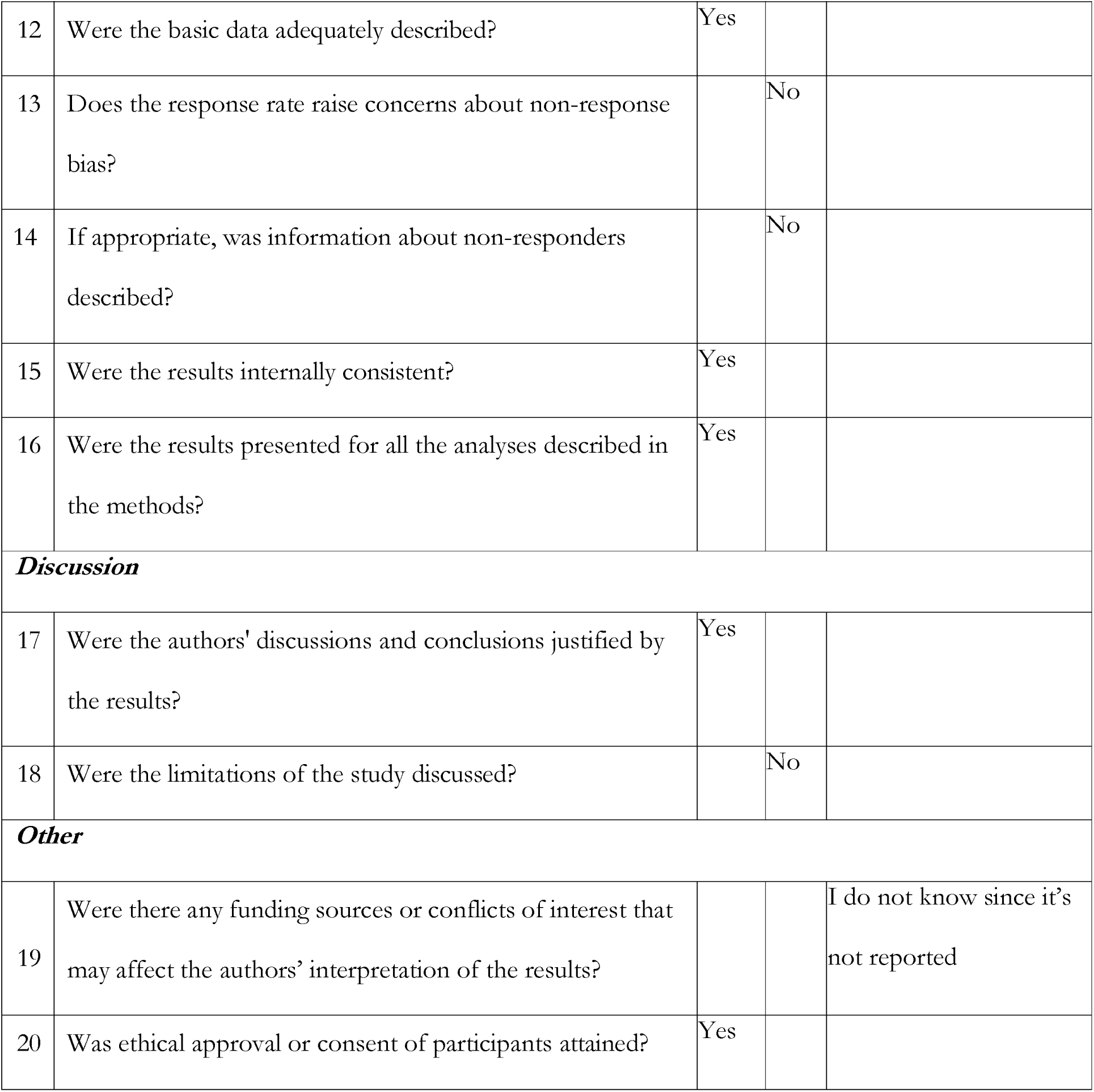

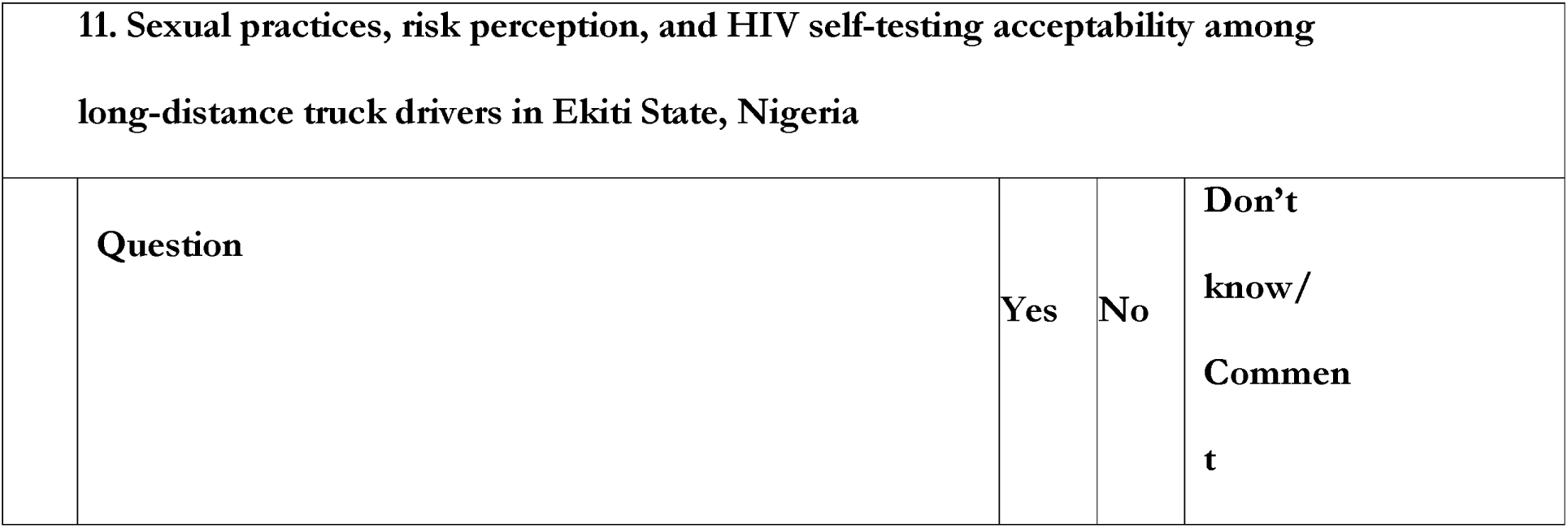
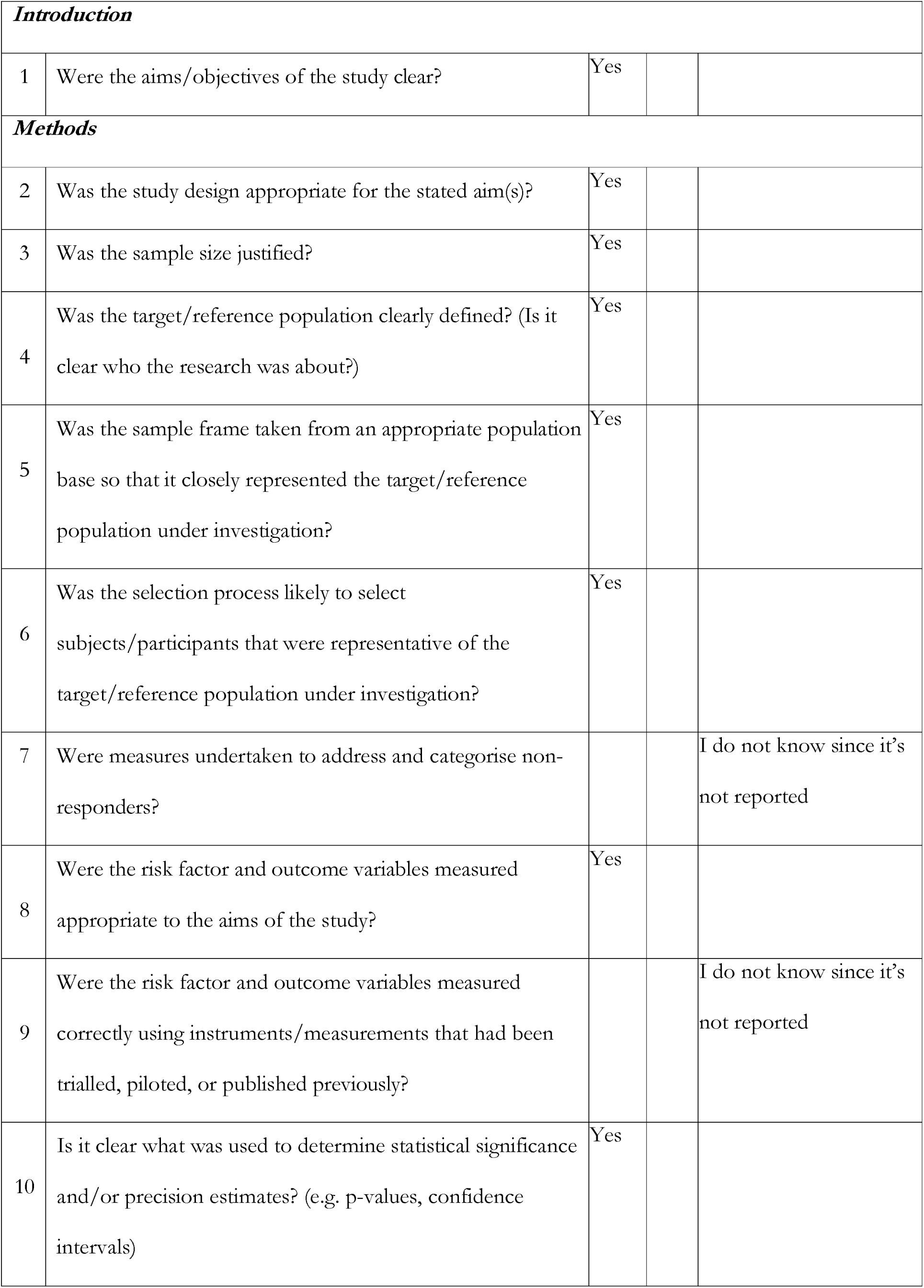
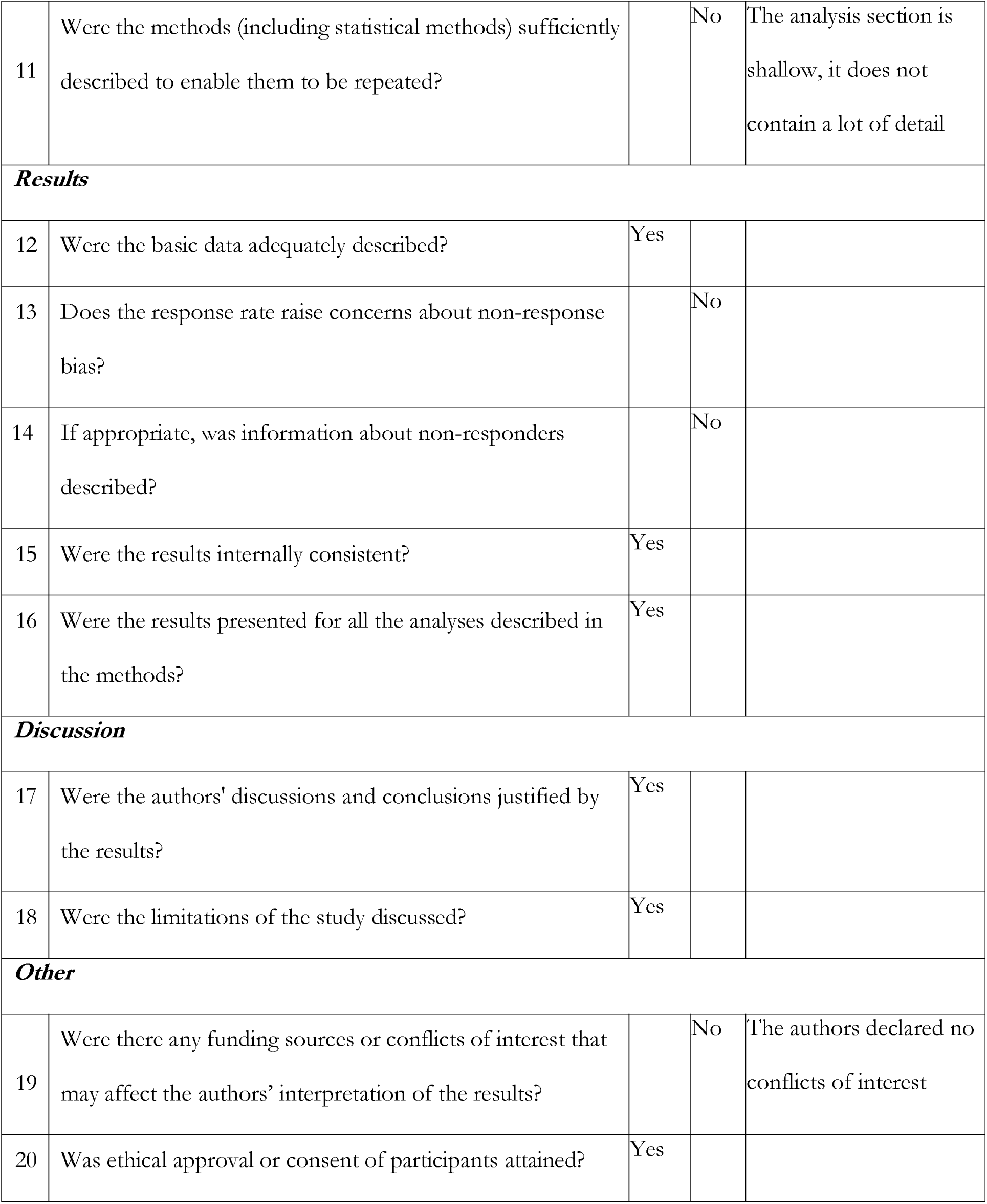

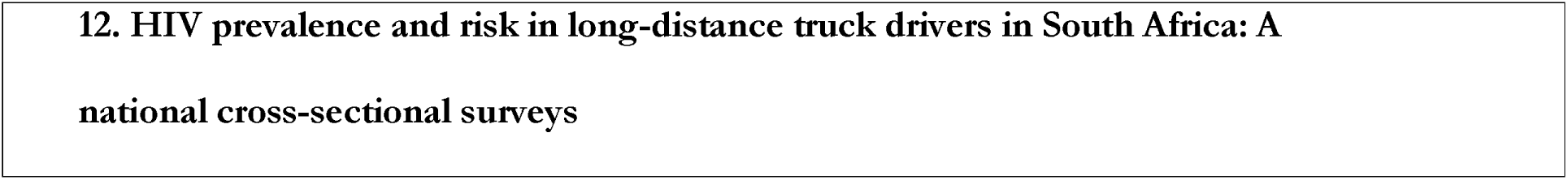
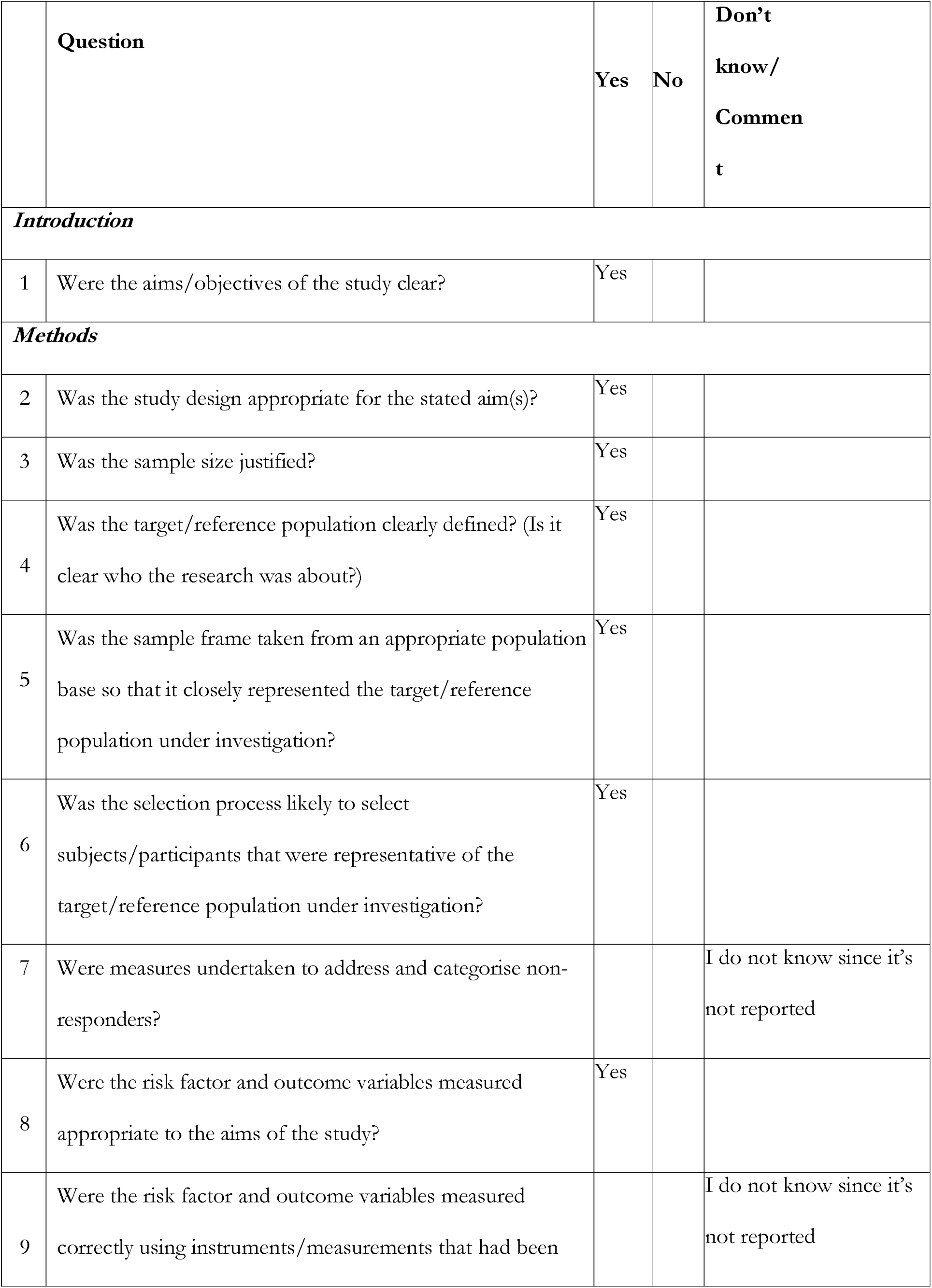
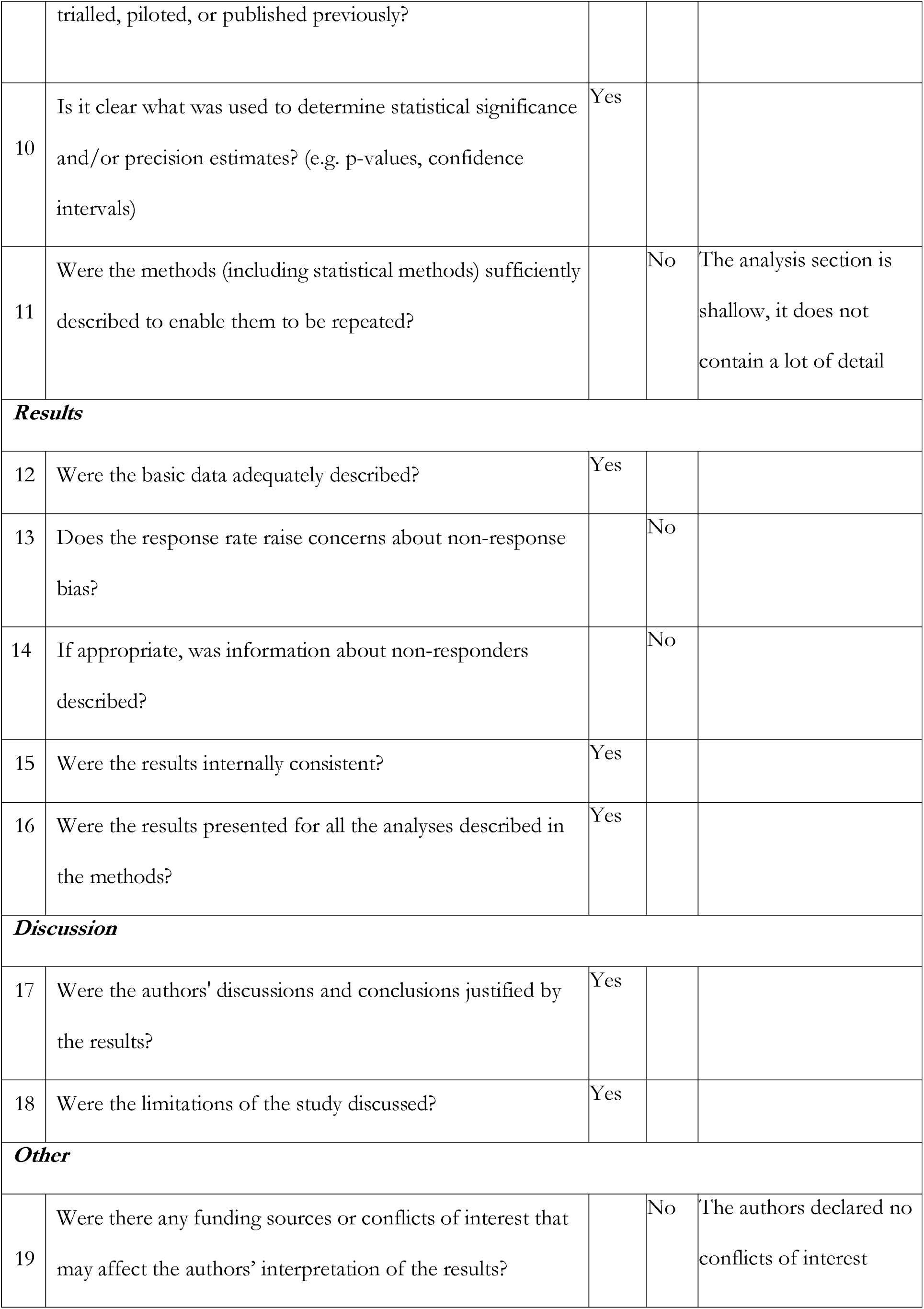
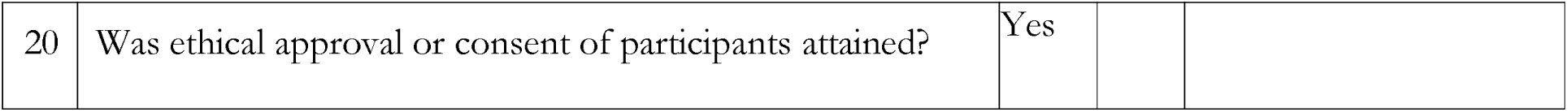

**12. HIV prevalence and risk in long-distance truck drivers in South Africa: A national cross-sectional surveys**
|  | Question | Yes | No | Don't know/<br>Comment |
| --- | --- | --- | --- | --- |
| **Introduction** |  |  |  |  |
| 1 | Were the aims/objectives of the study clear? | Yes |  |  |
| **Methods** |  |  |  |  |
| 2 | Was the study design appropriate for the stated aim(s)? | Yes |  |  |
| 3 | Was the sample size justified? | Yes |  |  |
| 4 | Was the target/reference population clearly defined? (Is it clear who the research was about?) | Yes |  |  |
| 5 | Was the sample frame taken from an appropriate population base so that it closely represented the target/reference population under investigation? | Yes |  |  |
| 6 | Was the selection process likely to select subjects/participants that were representative of the target/reference population under investigation? | Yes |  |  |
| 7 | Were measures undertaken to address and categorise non-responders? |  |  | I do not know since it's not reported |
| 8 | Were the risk factor and outcome variables measured appropriate to the aims of the study? | Yes |  |  |
| 9 | Were the risk factor and outcome variables measured correctly using instruments/measurements that had been trialled, piloted, or published previously? |  |  | I do not know since it's not reported |
| 10 | Is it clear what was used to determine statistical significance and/or precision estimates? (e.g. p-values, confidence intervals) | Yes |  |  |
| 11 | Were the methods (including statistical methods) sufficiently described to enable them to be repeated? |  | No | The analysis section is shallow, it does not contain a lot of detail |
| **Results** |  |  |  |  |
| 12 | Were the basic data adequately described? | Yes |  |  |
| 13 | Does the response rate raise concerns about non-response bias? |  | No |  |
| 14 | If appropriate, was information about non-responders described? |  | No |  |
| 15 | Were the results internally consistent? | Yes |  |  |
| 16 | Were the results presented for all the analyses described in the methods? | Yes |  |  |
| **Discussion** |  |  |  |  |
| 17 | Were the authors' discussions and conclusions justified by the results? | Yes |  |  |
| 18 | Were the limitations of the study discussed? | Yes |  |  |
| **Other** |  |  |  |  |
| 19 | Were there any funding sources or conflicts of interest that may affect the authors' interpretation of the results? |  | No | The authors declared no conflicts of interest |
| 20 | Was ethical approval or consent of participants attained? | Yes |  |  |

| **13. Cross-sectional study of the health of southern African truck drivers** |  |  |  |  |
| --- | --- | --- | --- | --- |
|  | **Question** | **Yes** | **No** | **Don't know/Comment** |
| ***Introduction*** |  |  |  |  |
| 1 | Were the aims/objectives of the study clear? | Yes |  |  |
| ***Methods*** |  |  |  |  |
| 2 | Was the study design appropriate for the stated aim(s)? | Yes |  |  |
| 3 | Was the sample size justified? |  | No | The study did not mention how they calculated the sample size |
| 4 | Was the target/reference population clearly defined? (Is it clear who the research was about?) | Yes |  |  |
| 5 | Was the sample frame taken from an appropriate population base so that it closely represented the target/reference population under investigation? | Yes |  |  |
| 6 | Was the selection process likely to select subjects/participants that were representative of the target/reference population under investigation? | Yes |  |  |
| 7 | Were measures undertaken to address and categorise non-responders? |  |  | I do not know since it's not reported |
| 8 | Were the risk factor and outcome variables measured appropriate to the aims of the study? | Yes |  |  |
| 9 | Were the risk factor and outcome variables measured correctly using instruments/measurements that had been trialled, piloted, or published previously? |  |  | I do not know since it's not reported |
| 10 | Is it clear what was used to determine statistical significance and/or precision estimates? (e.g. p-values, confidence intervals) |  | No | No statistical tests were conducted |
| 11 | Were the methods (including statistical methods) sufficiently described to enable them to be repeated? | Yes |  |  |
| **Results** |  |  |  |  |
| 12 | Were the basic data adequately described? | Yes |  |  |
| 13 | Does the response rate raise concerns about non-response bias? | Yes |  | Difficulty recruiting truckers |
| 14 | If appropriate, was information about non-responders described? |  | No |  |
| 15 | Were the results internally consistent? | Yes |  |  |
| 16 | Were the results presented for all the analyses described in the methods? | Yes |  |  |
| **Discussion** |  |  |  |  |
| 17 | Were the authors' discussions and conclusions justified by the results? | Yes |  |  |
| 18 | Were the limitations of the study discussed? | Yes |  |  |
| **Other** |  |  |  |  |
| 19 | Were there any funding sources or conflicts of interest that may affect the authors' interpretation of the results? |  | No | The authors declared no conflicts of interest |
| 20 | Was ethical approval or consent of participants attained? | Yes |  |  |

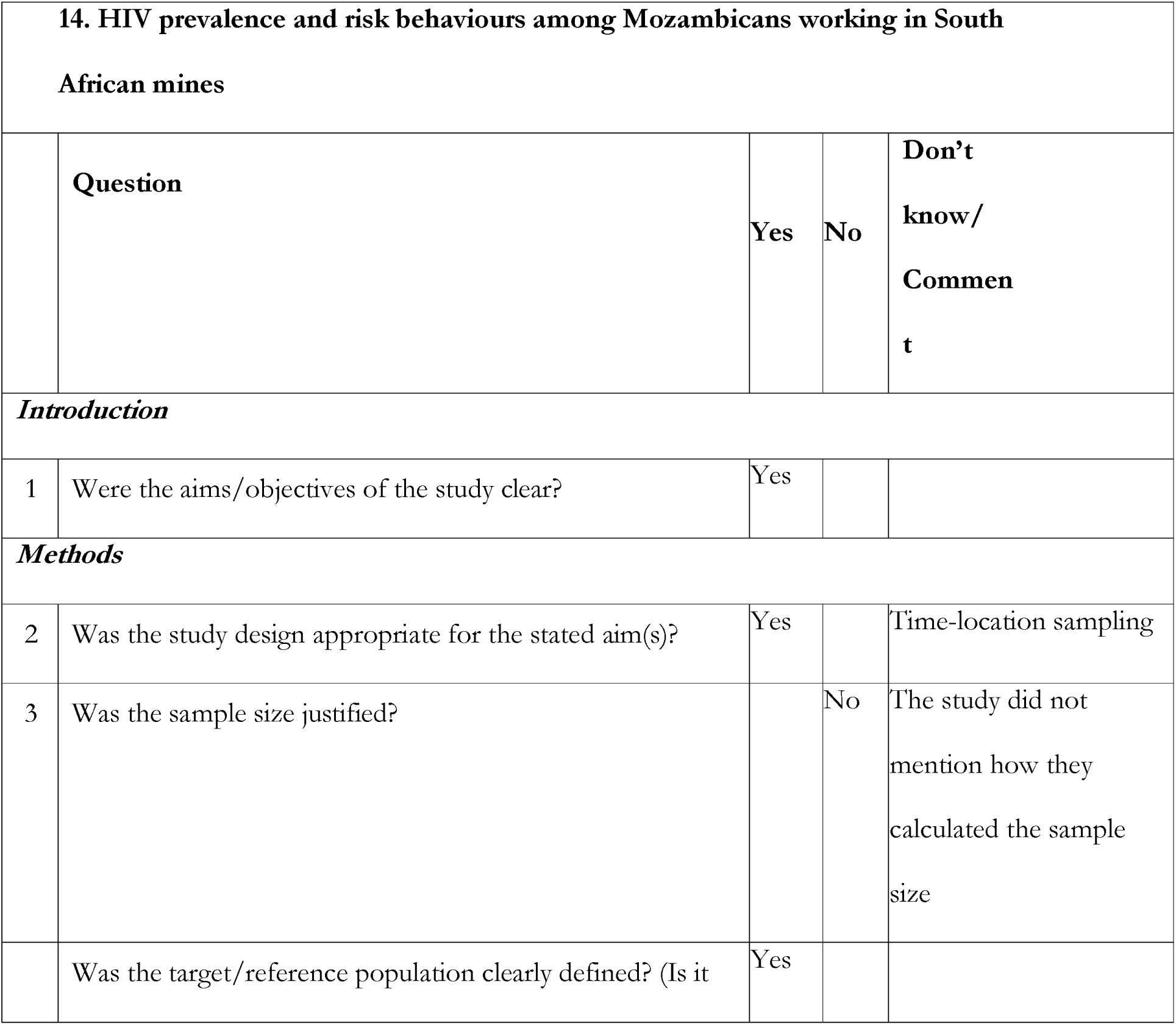
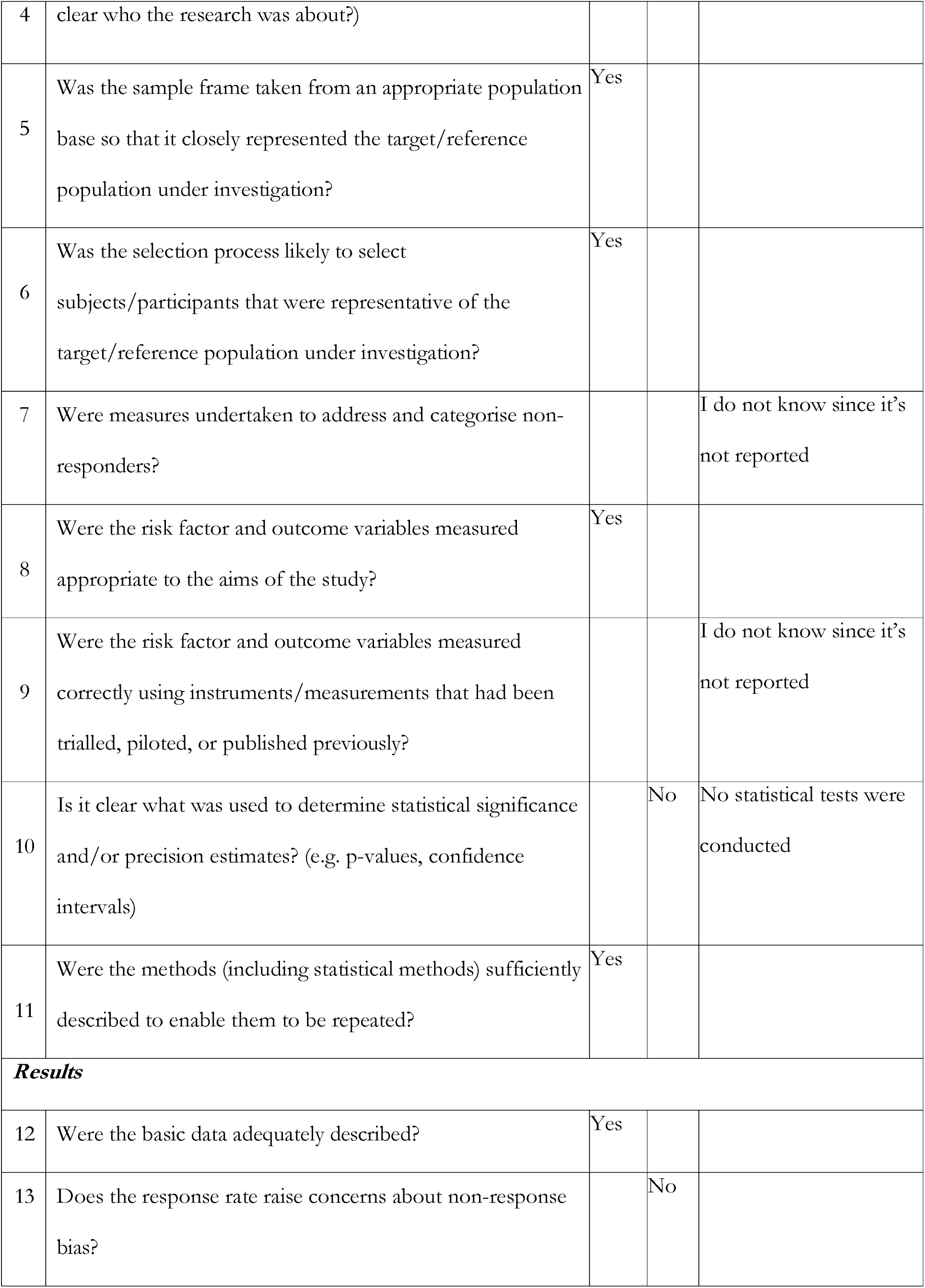
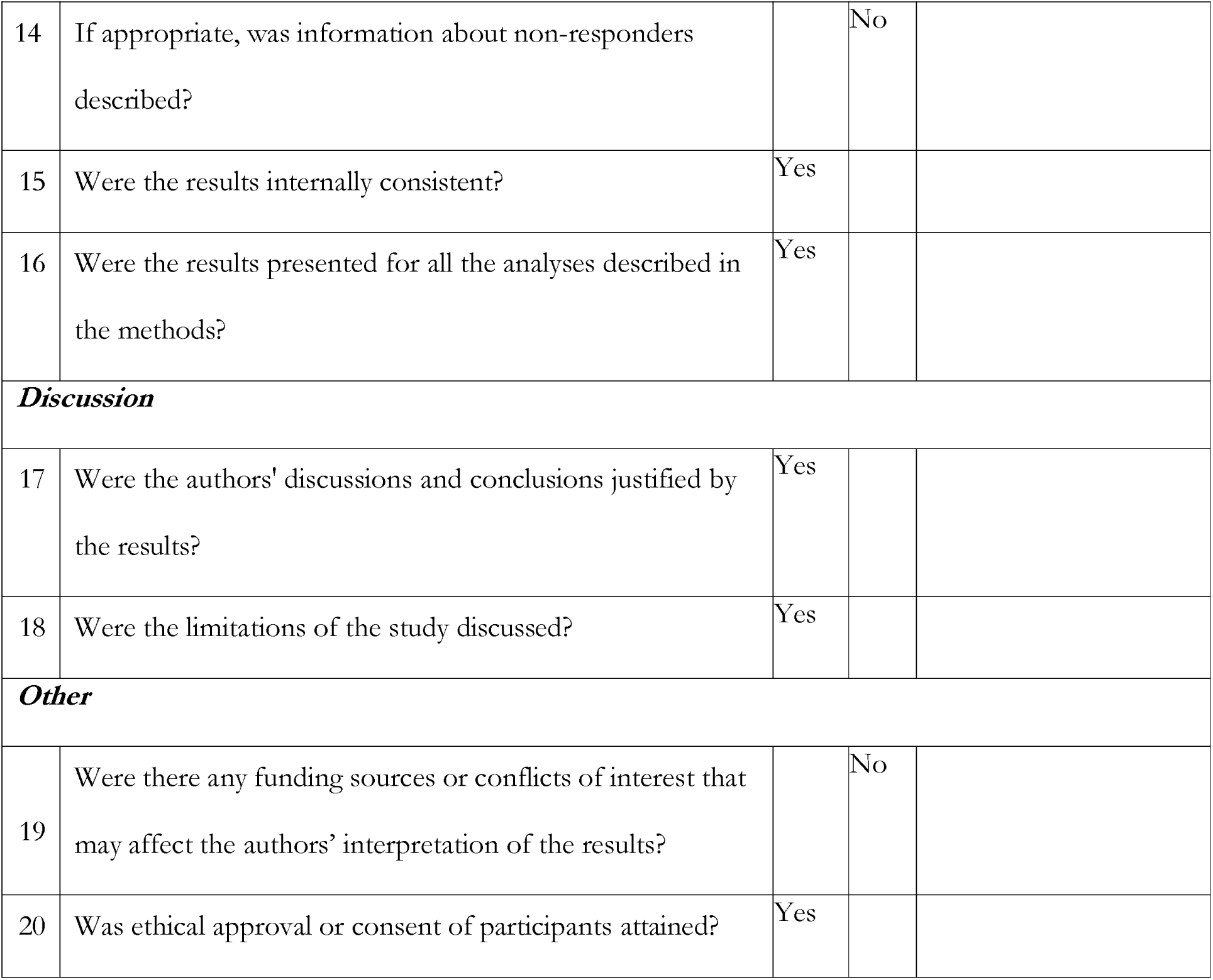

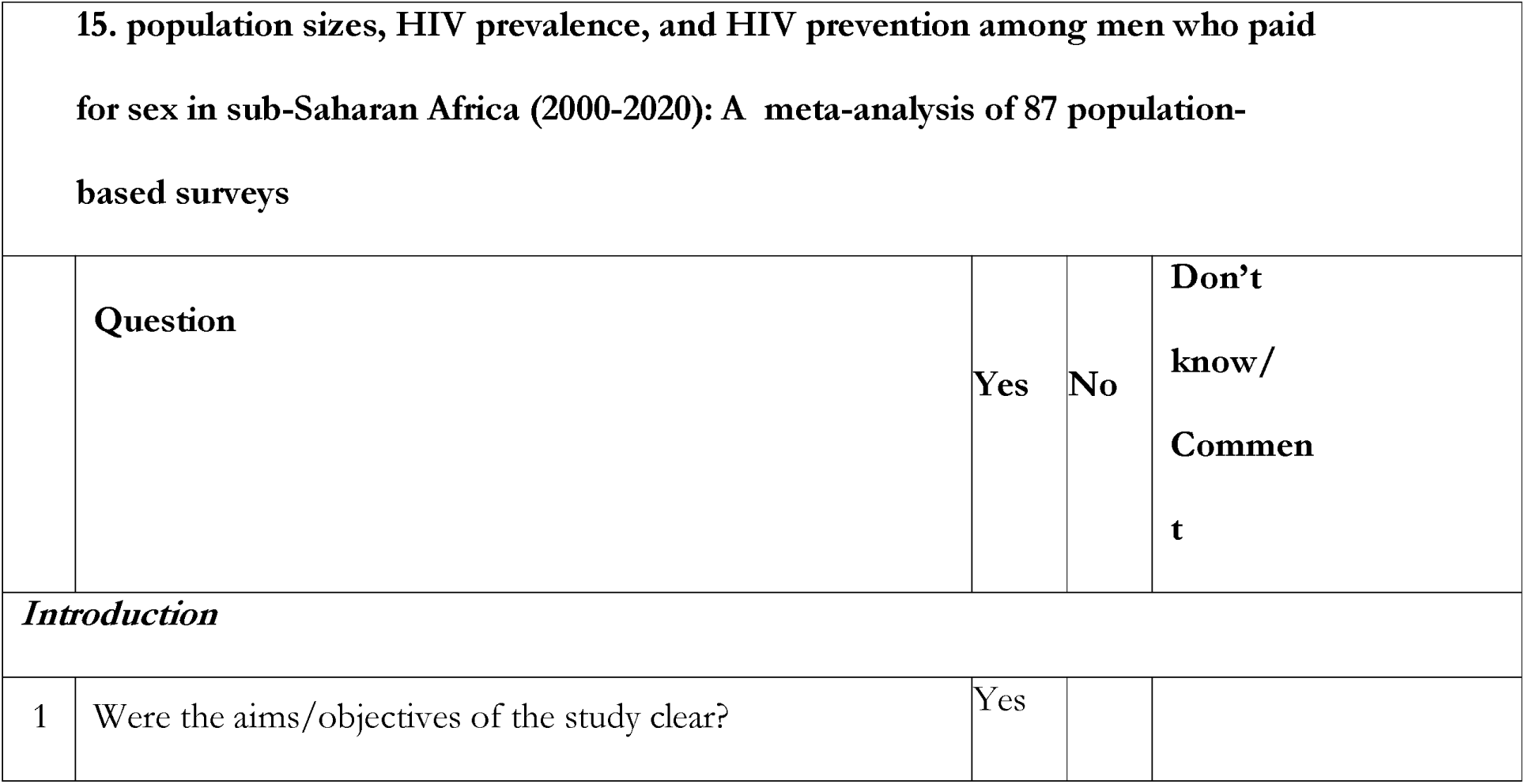
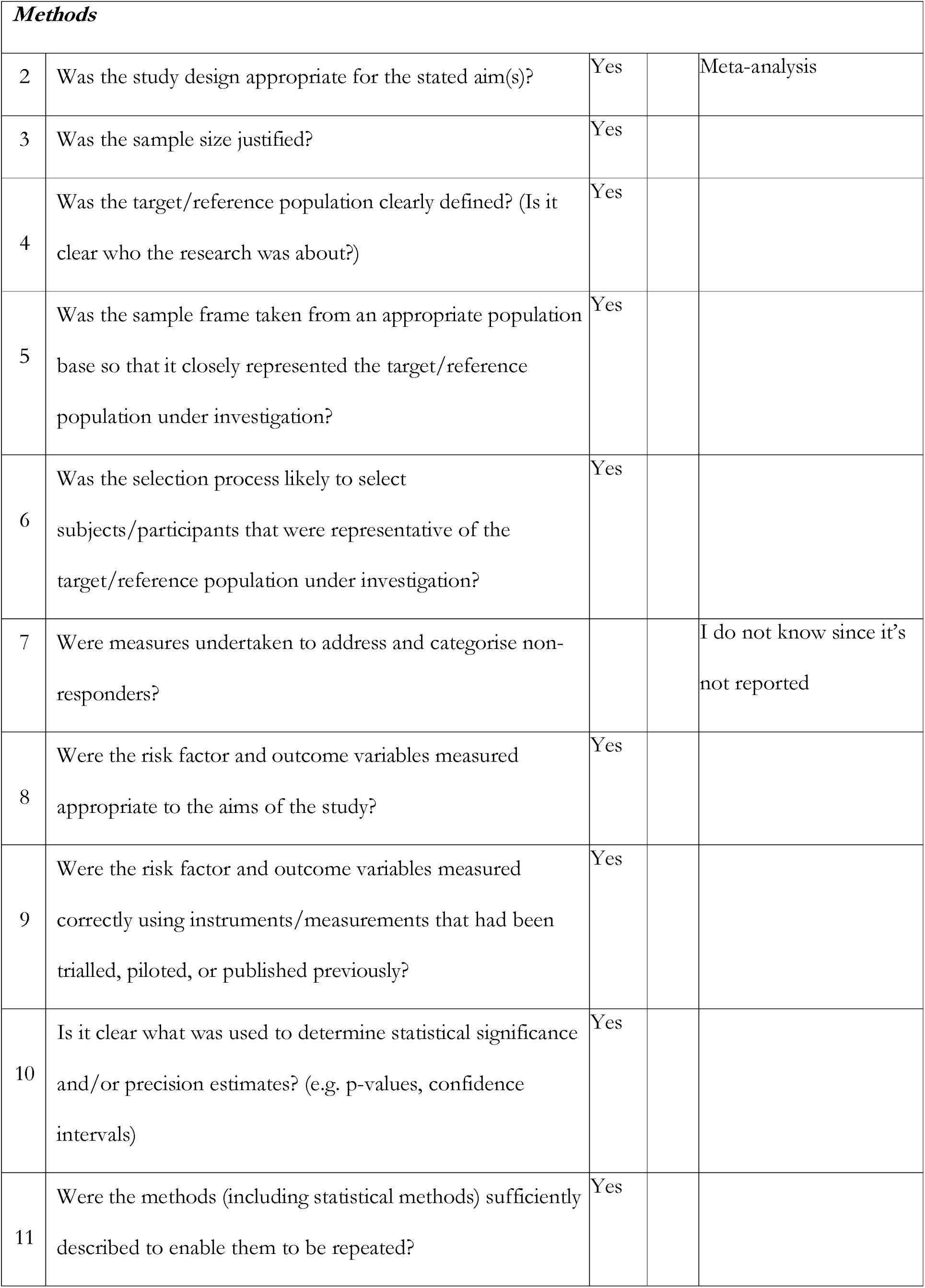
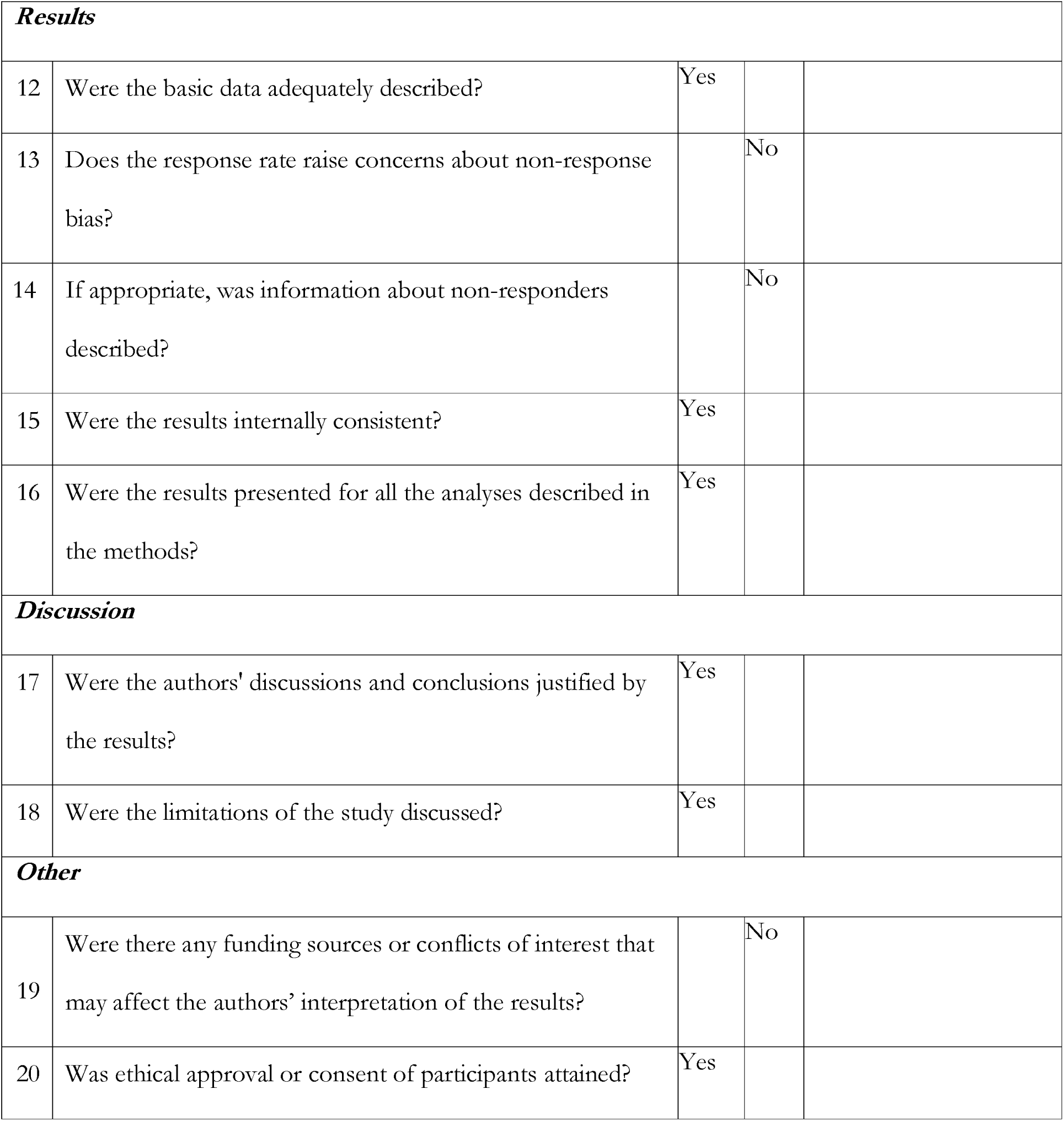

## S2 APPENDIX EXCLUDED STUDIES AND REASONS FOR EXCLUSION

S2 APPENDIX: EXCLUDED STUDIES AND REASONS FOR EXCLUSION
|  | Author | Reasons | Summary or description of the content |
| --- | --- | --- | --- |
| 1. | Assessing the Potential Impact of Disruptions Due to COVID-19 on HIV Among Key and Lower-Risk Populations in the Largest Cities of Cameroon and Benin | No data on having sex with a FSW | The study did not investigate whether the men had sex with FSW or not. |
| 2. | Differences in risky sexual behaviors and HIV prevalence of circumcised and uncircumcised men in Uganda: evidence from a 2011 cross-sectional national survey(66) | No data on having sex with a FSW | The study investigated sexual risk behaviours including transactional sex |
| 3. | Hazardous alcohol consumption and alcohol-related problems are associated with unknown and HIV-positive status in fishing communities in Uganda(67) | No data on having sex with a FSW | The study was assessed among different populations but did not investigate having sex with FSW among men recruited in the study |
| 4. | HIV risk perception and behavior among medically and traditionally circumcised males in South Africa(68) | No data on having sex with a FSW | • The study did not investigate whether the men had sex with FSW or not so it did not meet the inclusion criteria |
| 5. | Voluntary medical male circumcision and sexual practices among sexually active circumcised men in Mzuzu, Malawi: a cross-sectional study | No data on having sex with a FSW | • The study investigated transactional sex which is different from paying FSW for sex |
| 6. | An assessment of sexual risk behaviours among circumcised and uncircumcised men before and after the implementation of the safe male circumcision programme in Botswana(69) | No data on having sex with a FSW | • The study did not investigate having sex with FSW or not among the men recruited |
| 7. | Consistent condom use among men with non-marital partners in four sub-Saharan African countries(70) | No data on having sex with a FSW | • The study assessed the influence of marital status on condom use among men with casual partners. The article did not report about the men engaging female sex workers. |
| 8. | The contribution of emotional partners to sexual risk taking and violence among female sex workers in Mombasa, Kenya: a cohort study(71) | Ineligible population | • The study was conducted among FSW |
| 9. | HIV prevalence and risk behavior among male and female adults screened for enrolment into a vaccine preparedness study in Maputo, Mozambique(72) | Ineligible population | The study was conducted among men and women in the general community and did not report about men having sex with FSW. |
| 10. | Mobility is Associated with Higher-risk Sexual Partnerships Among Both Men and Women in Co-resident Couples in Rural Kenya and Uganda: A Longitudinal Cohort Study(73) | No data on having sex with a FSW | The study did not investigate having sex with female sex workers |
| 11. | Populations at increased risk for HIV infection in Kenya: results from a national population-based household survey, 2012(74) | No data on having sex with a FSW | The article reported about transactional sex with FSW which is different from sex work |
| 12. | Sexual risk behaviors following circumcision among HIV-positive men in Rakai, Uganda(75) | No data on having sex with a FSW | The study assessed sexual risk behaviours among HIV-positive men and did not include sex work engagement in this<br>population |
| 13. | Work-related violence and inconsistent condom use with non-paying partners among female sex workers in Adama City, Ethiopia(76) | Ineligible population | The study was conducted among female sex workers |
| 14. | Effects of a Short Message Service (SMS) Intervention on Reduction of HIV Risk Behaviours and Improving HIV Testing Rates Among Populations located near Roadside Wellness Clinics: A Cluster Randomised Controlled Trial in South Africa, Zimbabwe and Mozambique(77) | No data on having sex with a FSW | The study was conducted among truck drivers, sex workers, and community residents but did not report on any engagements between female sex workers and other populations. |
| 15. | Alcohol Misuse and Illicit Drug Use Among Occupational Groups at High Risk of HIV in Sub-Saharan Africa: A Systematic Review(78) | Ineligible study design | The article was a systematic review |
| 16. | A comparison of sexual risk behaviours and HIV seroprevalence among circumcised and | No data on having sex with a FSW | The study did not report on whether the men had sex with female sex workers or |
|  | uncircumcised men before and after implementation of the safe male circumcision programme in Uganda(79) |  | not. |
| 17. | Consistent condom use among men who pay for sex in sub-Saharan Africa: Empirical evidence from Demographic and Health Surveys(80) | Ineligible study design | The study used Demographic and Health Surveys that were also used in a meta-analysis conducted by Hodgins(81). |
| 18. | Factors Associated with Variations in Population HIV Prevalence across West Africa: Findings from an Ecological Analysis(82) | Ineligible study design | The study used data from Demographic and Health Surveys that were also used in the meta-analysis conducted by Hodgins(81) among 87 population based surveys. |
| 19. | Finding hidden HIV clusters to support geographic-oriented HIV interventions in Kenya(83) | Ineligible study design | This was a population-based study (KAIS) which was also reported in the meta-analysis by Hodgins(29) |
| 20. | Gender role conflict and sexual health and relationship practices amongst young men living in urban informal settlements in South Africa(84) | Ineligible population | The study was conducted among men and women in the general population |
| 21. | HIV epidemiology among female sex workers and their clients in the Middle East and North Africa: systematic review, meta-analyses, and meta-regressions(85) | Ineligible study design | The study was a systematic review and thus not primary research. |
| 22. | Khat chewing and risky sexual behavior in Sub-Saharan Africa: a systematic review protocol(86) | Ineligible study design | The study was a systematic review protocol and thus not primary research. |
| 23. | Social venues and HIV vulnerability of young venue workers: a cluster analysis of data from a national-level cross-sectional survey of social venues in Uganda(87) | Ineligible study design | This was a secondary analysis of data from a larger cross-sectional survey of social venues in 25 districts of Uganda. |
| 24. | Socioeconomic factors impact the risk of HIV acquisition in the township population of South Africa: A Bayesian analysis(88) | Ineligible study design | In this study, the authors conducted a secondary data analysis |
| 25. | Traditional Male Circumcision is Associated with Sexual Risk Behaviors in Sub-Saharan Countries | No data on having sex with a FSW | The study assessed the association between three sexual risk behaviors condom use |
|  | Prioritized for Male Circumcision(89) |  | with non-cohabiting partners, number of non-cohabiting partners, and partner type—and circumcision status but did not report about engaging FSW services among these men |
| 26. | Costing analysis of an SMS-based intervention to promote HIV self-testing amongst truckers and sex workers in Kenya(90) | No data on having sex with a FSW | • The study estimated the costs of an intervention to promote HIV testing among truck drivers and did not report on their engagement with female sex workers |
| 27. | Epidemiology of HIV among female sex workers, their clients, men who have sex with men, and people who inject drugs in West and Central Africa(91) | Ineligible study design | • The article is a systematic review of HIV literature among key populations in WCA conducted since the onset of the HIV epidemic. |
| 28. | Evaluating the prevalence and patterns of sexual risk behavior among undergraduate university students in Ethiopia: implication for psychosocial intervention(92) | Results different from what the study design talks about | • There is a difference between what the study seeks to achieve and the findings of the study |
| 29. | HIV and Syphilis Prevalence and Associated Risks in the Cameroonian Armed Forces(93) | No access to full-text | There is no access to full-text for this study |
| 30. | HIV Testing Approaches to Optimize Prevention and Treatment for Key and Priority Populations in Malawi(94) | Ineligible study design | The data used for analysis was extracted from program data routinely collected for the LINKAGES project |
| 31. | HIV-related stigma and its association with HIV transmission risk behaviors among boda boda motorcyclists in Mbarara municipality, southwestern Uganda(95) | No access to full text | There was no access to full-text for the article. |
| 32. | HIV/AIDS risk perception and behavior of college students of the Metekel Zone, Benishangul Gumuz regional state, Ethiopia(96) | No access to full-text | There was no access to full-text for the article. |
| 33. | Predictors for HIV testing among Chinese workers in infrastructure construction enterprises in Kenya(97) | No data on having sex with a FSW | The study did not investigate whether the men recruited in the study had sex with FSW or not which was an important |

|  |  |  | inclusion criteria |
| --- | --- | --- | --- |
| 34. | Predictors of HIV Among 1 Million Clients in High-Risk Male Populations in Tanzania(98) | Ineligible study design | This article used secondary data from a cross-sectional study nested within a large-scale community-based HIV prevention program |
| 35. | Role Of Emigration In The Spread Of HIV Infection In The Region Of Kayes, Mali(99) | Inapplicable language- French | The article was written in French and I could not translate it. |
| 36. | Socio-demographic contributors to health and safety of mine workers in South Africa(100) | No data on having sex with a FSW | The study assessed the relationship between socio-demographic factors and health, safety, and wellness among mine workers. There was no investigation of having sex with female sex workers or not. |
| 37. | Transactional sex and HIV risks - evidence from a cross-sectional national survey among young people in Uganda(101) | No data on having sex with a FSW | The study assessed transactional sex which is different from paying FSW for sex |
| 38. | Uptake and retention on HIV pre-exposure | Ineligible study | The study used secondary data from |
|  | prophylaxis among key and priority populations in South-Central Uganda(102) | design | registers of the PrEP programme. |
| 39. | What's in a name? A mixed method study on how young women who sell sex characterize male partners and their use of condoms(103) | Inapplicable population | • The study was conducted among female sex workers to characterize male partners and their use of condoms. The study did not investigate sex work among men. |
| 40. | Willingness to use short-term oral pre-exposure prophylaxis (PrEP) by migrant miners and female partners of migrant miners in Mozambique(104) | No access to full-text | • There is no access to the full text of the article<br>• The article does not report on paying sex worker for sex |
| 41. | Alcohol use and its association with sexual risk behaviors in the Armed Forces of the Republic of the Congo(32) | Ineligible study design | • A secondary analysis of data collected from a 2014 seroprevalence and behavioral epidemiology risk survey was performed |
| 42. | Knowledge, attitudes & barriers to condom use among female sex workers and truck drivers in Uganda: a mixed-methods study(105) | No data on having sex with a FSW | • The study did not investigate whether there was FSW engagement<br>• The study also employed mixed methods<br>approach |
| 43. | HIV/AIDS prevention practices among military personnel in Northwest Ethiopia(106) | No data on having sex with a FSW | The study did not investigate whether the military personnel had sex with FSW or not |
| 44. | The association between client type and condom use with steady and unsteady partners among persons seeking HIV testing and counseling services in Kenya | No data on having sex with a FSW | The study did not investigate FSW engagement among the population surveyed<br>Assessments were made on the general population, truck drivers, and FSW |
| 45. | Consistent Condom Use during Casual Sex among Long-Truck Drivers in Togo(107) | No data on having sex with a FSW | The study assessed factors associated with casual sex among LDTD which is different from sex with FSW |
| 46. | Comportment of heavy goods vehicle drivers in HIV spread along settlements around Kaduna: Kano road transport corridor in Nigeria(108) | No data on having sex with a FSW | The study did not investigate about having sex with FSW which is an important inclusion criteria |
| 47. | Left behind?: male clients of female sex workers in Zambia | Ineligible study design | This was a Demographic and Health Survey (DHS) which was also part of the meta-analysis. |
| 48. | Factors associated with buying sex and the knowledge that condoms prevent HIV among longdistance truck drivers at Kazungula weighbridge terminal, Chobe District, Botswana(33) | Duplicate article | • The paper reports the same results as reported in another article but with a different title. |
| 49. | Condom Use Behaviors and Correlates of Use in the Botswana Defence Force(109) | No data on having sex with an FSW | • The inclusion criteria for this scoping review specifically require the presence of having sex with FSW among the subgroups of men. |

